# Vaccine Adverse Event Reporting System (VAERS): Evaluation of 31 Years of Reports and Pandemics’ Impact

**DOI:** 10.1101/2022.03.23.22272819

**Authors:** Ohoud A. Almadani, Thamir M. Alshammari

## Abstract

**Background:** Vaccine adverse event reporting system (VAERS) was established in the United States (U.S.) as an early warning system with a main purpose of collecting postmarketing adverse events following immunizations (AEFIs) reports to monitor the vaccine safety and to mitigate the risks from vaccines. During the coronavirus diseases 2019 (COVID-19) pandemic, VAERS got more attention as its important role in monitoring the safety of the vaccines. Thus, the aim of this study was to investigate VAERS patterns, reported AEFIs, adverse events of special interest (AESIs), impact of different pandemics since its inception, and surveillance rate of serious vs nonserious AEFIs.

**Methods:** This was an observational study using VARES data from 2/7/1990 to 12/11/2021. Patterns of reports over years were first described, followed by a comparison of reports statistics per year. Furthermore, a comparison of incidents (death, ER visits, etc.) statistics over years, in addition to statistics of each vaccine were calculated. Moreover, each incident’s statistics for each vaccine were calculated and top vaccines were reported. Finally, survival analysis utilizing cox regression was done, in addition to AESIs distribution stratified by age groups and gender. All analyses were conducted using R (Version 1.4.1717) and Excel for Microsoft 365.

**Results:** There were 1,396,280 domestic and 346,210 non-domestic reports during 1990-2021, including 228 vaccines. For both domestic and non-domestic reports, year of 2021 had the highest reporting rate (48.52% and 70.33%), in addition a notable changes in AEFIs patterns were recorded during 1991, 1998, 2000, 2006, 2009, 2011, and 2017. AEFIs were as follow: deaths (1.00% and 4.08%), ER or doctor visits (13.37% and 2.27%), hospitalizations (5.84% and 27.78%), lethal threat (1.42% and 4.38%), and disabilities (1.4% and 7.96%). Pyrexia was the top reported symptom during the past 31 years, except for 2021 where headache was the top one. COVID-19 vaccines namely Moderna, Pfizer-Biontech, and Janssen were the top 3 reported vaccines with headache, pyrexia, and fatigue as the top associated AEFIs. Followed by Zoster, Seasonal Influenza, Pneumococcal, and Human papillomavirus vaccines. Myocarditis or Pericarditis were the top reported AESIs (26.95% and 25.84%). Male (HR:1.15, 1.14 - 1.16) for domestic (HR:1.23, 1.22 -1.25) for nondomestic have a higher probability of having serious AEFIs. In addition, age group ≤ 5 years old in domestic and >84 years old in nondomestic have a higher probability of having AEFIs compared to other age groups.

**Conclusions:** The large data available in VARES make it a useful tool for detecting and monitoring vaccine AEFIs. However, its usability relies on understating the limitations of this surveillance system, the impact of governmental regulations, availability of vaccines, and public health recommendations on the reporting rate.

## I. Introduction

The Council for International Organizations of Medical Sciences (CIOMS) defined an adverse event following immunization (AEFI) as “any untoward medical occurrence which follows immunization, and which does not necessarily have a causal relationship with the usage of the vaccine. AEFI may be any unfavorable or unintended sign, abnormal laboratory finding, symptom or disease” ^1–3^.

To identify AEFIs, vaccine testing for safety is conducted in laboratories on animals and humans in clinical trials, similar to medications’ approval process ^2,4–8^. Three phases of clinical trials are required to determine the tested vaccine’s effectiveness (immunogenicity) and safety (including adverse events) ^2,4–7,9^.

The United States Food and Drug Administration (USFDA) requires this premarketing evaluation process before granting any vaccine licensure ^2,8,9^. Such evaluations effectively detect the most common AEFIs of tested vaccines ^2,10^. However, some AEFIs cannot be detected easily, especially rare ones, either due to the sample size or duration of premarketing trials ^8,10,11^.

The Vaccine Adverse Event Reporting System (VAERS) is an early warning system that the USFDA and Centers for Disease Control and Prevention (CDC) jointly established in 1990. The main purpose of this system is to collect postmarketing AEFIs reports ^12^.

The VAERS received more attention following the coronavirus disease 2019 (COVID-19). Majority of these vaccines were approved under emergency use authorization (EUA), which require health care providers to record certain AEFIs in the VAERS, including death ^13–15^.

Since the establishment of the VAERS, no study has been conducted to evaluate it comprehensively. Therefore, we aimed to provide a descriptive analysis of VAERS data from its inception to 2021, focusing on pattern analysis of reported vaccines, reported AEFIs, reported admissions, and other related variables. We also aimed to describe the most frequently reported symptoms and vaccines that have been associated with each of them, and adverse events of special interest (AESIs) associated with COVID-19. Last, we aimed to calculate serious AEFIs survival probability.

## II. Materials and Methods

### A. Study Design

This was a retrospective observational study using the VAERS from the first available report in the data (July 2, 1990) to the last available report (November 12, 2021).

### B. Data Sources

This study includes data from VAERS, which is a passive AEFI reporting system that the USFDA and CDC established in 1990. These multidimensional data are reported by individual health care providers or the public when someone experiences an AEFI, without a confirmation of the AEFI’s real cause. Data available on the website is formatted as ready-to-download comma-separated value (CSV) files, which can be imported to the CDC WONDER online search tool. VAERS data are available as domestic and nondomestic data. More than 99% of nondomestic data is reported by vaccine manufacturers. Furthermore, these data are divided for each year into three files: data, symptoms, and vaccine information. Table 1 presents variables available in VAERS data that we have used in this paper. The code of federal regulations (CFR) defines a lethal threat as any adverse event or condition that puts the patient or subject in immediate danger of death. In addition, it defines disability as a long-term or considerable inability or disturbance of the subject’s ability to carry out normal life functions ^7^. Moreover, an AESI is a condition or event that occurs in certain people after vaccination and has the potential to be causally linked to a vaccine ^16–18^. AESIs we included in this study are anaphylaxis, appendicitis, bell’s palsy, deep vein thrombosis (DVT), disseminated intravascular coagulation, encephalomyelitis, Guillain-Barré syndrome, immune thrombocytopenia, myocardial infarction, myocarditis or pericarditis, narcolepsy, pulmonary embolism, non-hemorrhagic or hemorrhagic stroke, and transverse myelitis ^16–18^.

**Table 1:** VAERS data variables names and descriptions

| **Variable** | **Description** |
| --- | --- |
| **VAERS_ID** | VAERS identification number which is available in the data, symptoms, and vaccines files. |
| **RECVDATE** | Receiving data of the report, which has been converted to year. |
| **AGE_YRS** | Age in years. |
| **SEX** | Reported gender. |
| **DIED** | Patient died following vaccination. |
| **L_THREAT** | Patient has life-threatening illness any time following vaccination. |
| **ER_VISIT** | Patient visits ER any time following vaccination. |
| **HOSPITAL** | Patient been hospitalized any time following vaccination. |
| **HOSPDAYS** | Number of hospitalization days. |
| **DISABLE** | Patient has disability any time following vaccination. |
| **VAX_TYPE** | Administered vaccine type |
| **VAX_NAME** | Vaccination name |
| **SYMPTOM** | Adverse event as per MedDRA* |

\* MedDRA is an international medical terminology.

### C. Data reprocesses

VAERS data is multidimensional data, meaning one report may contain several symptoms or vaccines. First, for domestic data, we merged symptoms and vaccine files based on VAERS_ID. Then we separated reports containing multiple vaccines—because no clear information is available about which vaccine triggered those events—in a way that each vaccine reported in them presents the same event information. We followed the same steps with nondomestic files. Reprocessing completed using R studio (version 1.4.1717).

### D. Data analysis

We first described patterns of reports over the years by calculating the number of reports per year and then placing that number on a line chart. We then calculated a comparison of reports statistics per year as follows:

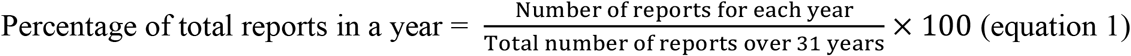

Furthermore, we compared incident (death, ER visits, etc.) rates in a year as follows:

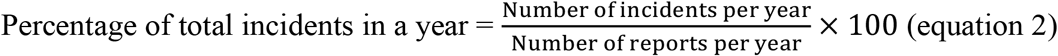

In addition, we calculated yearly statistics for each vaccine, and we then described the most common vaccine patterns using line charts.

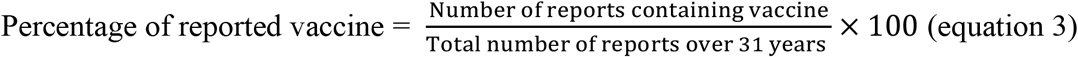

Moreover, we calculated the incident statistics (death, ER visits, etc.) for each vaccine, and we reported the most common vaccines.

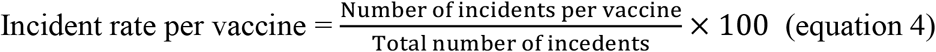

We made these calculations and created these charts using Excel for Microsoft 365. We excluded 2019 coronavirus disease (COVID-19) vaccines from the charts of the most common vaccine patterns due to their recent development.

Moreover, we performed a survival (time to event) analysis, were individuals followed up from the vaccination date up to the event date (onset of AEFIs), using survival package in R studio^19^. Based on FDA protocol recommendation, we restrict follow up to 365 days ^16,20^. All data in VAERS are for confirmed events, because of that we used non serious AEFIs – instead of no event - as a comparator to serious AEFIs to calculate the probability of survival ^19^.

Last, we calculated distribution of AESIs among all vaccines, and their distribution to COVID vaccines.

## III. Results

### A. Domestic data

There were 1 396 280 reports from 1990 through 2021. Reports included 228 vaccines. The

most reports were recorded in 2021 (n = 677 514, 48.52%), followed by 2020 (n = 50 205, 3.60%), 2018 (n = 49 137, 3.52%), 2019 (n = 48 443, 3.47%), and 2016 (n = 45 706, 3.27%).

Most of the samples were from female subjects (60.90%), and the subjects’ average age was 27.8 years (max = 119, min = <1 month). Supplementary figure 1 shows the number of domestic reports per year.

COVID-19 vaccines were the top 3 reported vaccines—Moderna (19.81%), Pfizer-Biontech (19.45%), and Janssen (4.46 %)—followed by Zoster (Shingrix; 4.10%), seasonal influenza (Fluzone; 2.88%), Pneumo (Pneumovax; 2.87%), Zoster live (Zostavax; 2.41%), HPV (Gardasil; 2.38%), Varicella (Varivax; 2.08%), and measles-mumps-rubella (MMR II; 1.96%). Supplementary table 2 presents the number of domestic and nondomestic reports per vaccine.

### Reported deaths

The total number of reported deaths is 13 926 (1.00%), with the most deaths reported in 1990 (3.62%), followed by 1991 (2.62%), 1994 (2.19%), 1993 (2.16%), and 1992 (2.04%). Reported deaths continuously decreased over the past 30 years except for 2021, when the percentage of reported deaths increased. Supplementary figure 2 shows pattern of reported death over the past 31 years.

The top vaccines in terms of reported deaths are Pfizer-Biontech (27.98%), Moderna (24.01%), Janssen (7.16%), seasonal influenza, no brand (3.24%), and Fluzone (2.91%). Supplementary figure 3 presents the top vaccines in terms of reported deaths, excluding COVID vaccines due to their recent addition. Supplementary table 3 presents the number of reported deaths per vaccine per year.

### Reported ER or doctor office visits

The total number of reported ER visits is 186 645 (13.37%), with the most ER visits occurring in 2007 (47.31%). Supplementary figure 4 shows the percentage of ER visits per year.

Fluzone (7.22%) is the top vaccine in terms of reported ER visits, followed by Gardasil (6.00%), Pneumovax (4.98%), Varivax (4.05%), and MMRII (4.03%). Supplementary figure 5 presents the top five vaccines in terms of reported ER visits. In addition, supplementary table 4 presents the number of domestic reported ER or doctor visits per vaccine per year.

### Reported hospitalizations

The number of reported hospitalizations is 81 538 (5.84%), with a continuous decrease in the hospitalization rates and the largest hospitalization rate occurring in 1990 (12.13%). Supplementary figure 6 shows pattern of domestic reported hospitalization over years.

The highest number of average hospitalization days (mean = 12.7 days) occurred in 2021, compared to an average of hospitalization days of 6.3 over the past 30 years. COVID vaccines are the top vaccines in terms of hospitalization—Pfizer-Biontech (28.09%), Moderna (20.34%), and Janssen (6.62%)—followed by seasonal influenza, no brand (2.31%), Fluzone (2.28%), and Gardasil (2.07%). Supplementary table 5 presents the number of reported hospitalizations per year with the average number of hospitalization days. In addition, supplementary figure 7 presents the top vaccines in terms of reported hospitalization.

### Reported lethal threats

The number of reported lethal threats is 19 872 (1.42%), with the most lethal threats occurring in 1999 (2.56%), followed by 2.03% in 1995 and 2.00% in 1994. Supplementary figure 8 presents the pattern of reported lethal threats over years.

COVID vaccines are the top vaccines in terms of lethal threats—Pfizer-Biontech (22.35%), Moderna (18.23%), and Janssen (6.04%)—followed by Fluzone (4.13%), Gardasil (3.76%), and seasonal influenza, no brand (2.21%). Supplementary figure 9 presents the top five vaccines in terms of reported lethal threats per years. In addition, supplementary table 6 presents the number of domestic reported lethal threats per vaccine per year.

### Reported disabilities

The total number of reported disabilities is 22 639 (1.4%), with the most frequently reported disabilities occurring in 1990 (3.95%), followed by 2002 (2.87%) and 2001 (2.81%). Supplementary figure 10 presents the pattern of reported disabilities over years.

Pfizer-Biontech is the top vaccine in terms of reported disabilities (20.04%), followed by Moderna (15.55%), Gardasil (5.35%), Janssen (4.58%), and Zostavax (4.29%). Supplementary figure 11 shows the top five vaccines in terms of reported disabilities. In addition, supplementary table 7 presents the number of domestic reported disabilities per vaccine per year.

### Most frequently reported symptoms over the years

The total number of reported symptoms is 3 449 887. Table 8 lists the top vaccines in terms of reported symptoms. From 1990 through 2000, pyrexia was the most frequently reported AEFI (8.41%), followed by injection site hypersensitivity (4.29%), rash (3.36%), injection site edema (3.14%), and vasodilatation (2.88%). However, from 2001 through 2010, erythema was the most frequently reported AEFI (7.16%), followed by pyrexia (4.59%), injection site pain (4.57%), injection site swelling (2.55%), and rash (2.16%). Finally, from 2011 through 2020, pyrexia was the most frequently reported AEFI (3.33%), followed by injection site pain (2.85%), injection site erythema (2.73%), incorrect product storage (2.15%), and injection site swelling (2.04%). Table 9 presents the most frequently reported AEFIs and the associated vaccines.

**Table 8:** Top vaccines in reported symptoms

| **Vaccine** | **%** | **Count of reported symptoms** |
| --- | --- | --- |
| **COVID19 (Moderna)** | 25.51% | 880006 |
| **COVID19 (Pfizer-Biontech)** | 25.05% | 864213 |
| **COVID19 (Janssen)** | 5.74% | 198136 |
| **Zoster (Shingrix)** | 5.28% | 182011 |
| **Influenza (Seasonal) (Fluzone)** | 3.57% | 123257 |
| **Pneumo (Pneumovax)** | 3.39% | 117097 |
| **HPV (Gardasil)** | 2.72% | 93746 |
| **Zoster live (Zostavax)** | 2.70% | 92977 |
| **Influenza (Seasonal) (no brand name)** | 1.85% | 63899 |
| **Measles + Mumps + Rubella (MMR II)** | 1.80% | 62228 |
| **Varicella (Varivax)** | 1.70% | 58695 |
| **Influenza (Seasonal) (Fluzone high-dose)** | 1.61% | 55405 |

**Table 9:**
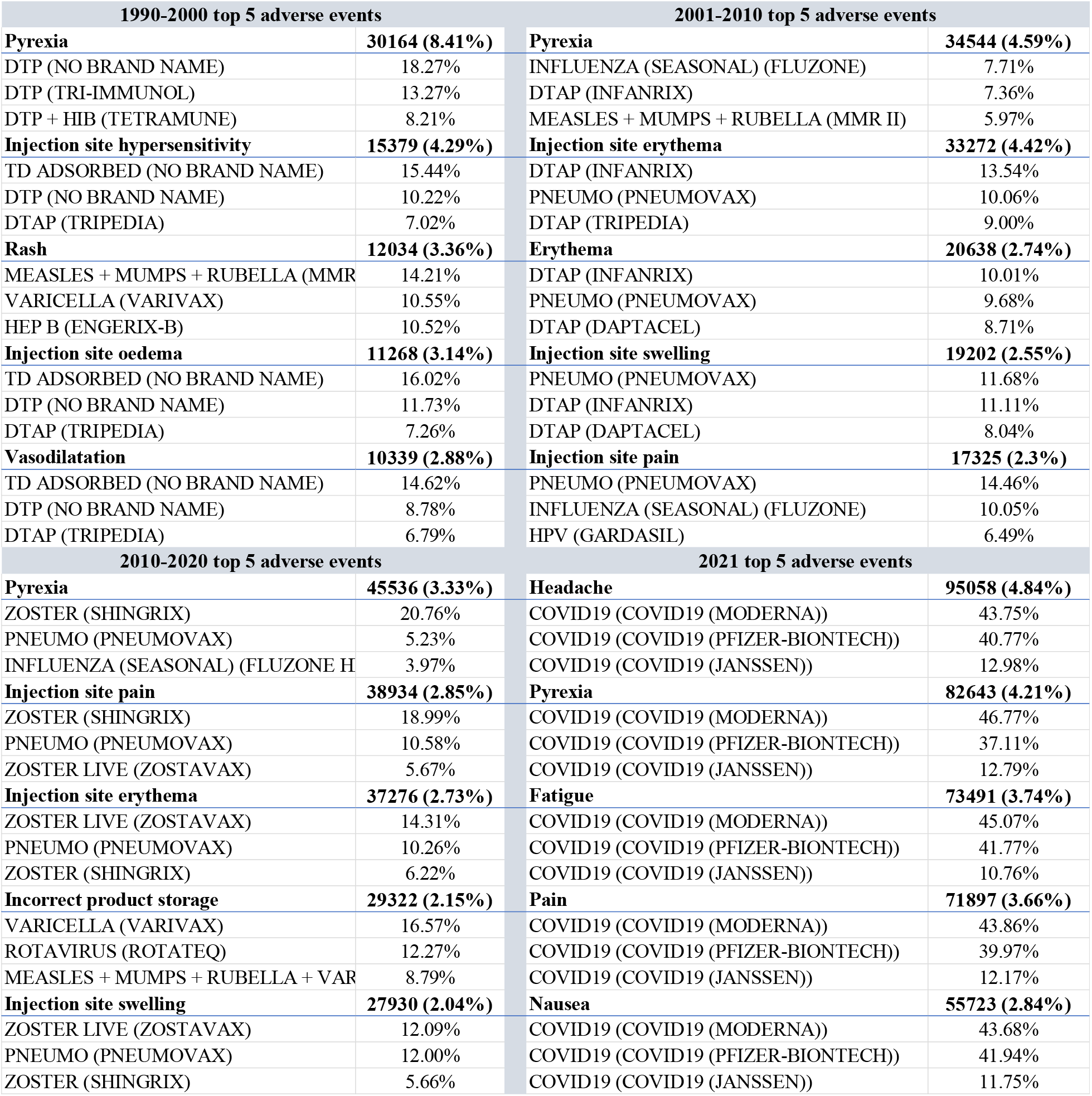
Top reported AEFI with top related vaccines

| 1990-2000 top 5 adverse events |  | 2001-2010 top 5 adverse events |  |
| --- | --- | --- | --- |
| **Pyrexia** | **30164 (8.41%)** | **Pyrexia** | **34544 (4.59%)** |
| DTP (NO BRAND NAME) | 18.27% | INFLUENZA (SEASONAL) (FLUZONE) | 7.71% |
| DTP (TRI-IMMUNOL) | 13.27% | DTAP (INFANRIX) | 7.36% |
| DTP + HIB (TETRAMUNE) | 8.21% | MEASLES + MUMPS + RUBELLA (MMR II) | 5.97% |
| **Injection site hypersensitivity** | **15379 (4.29%)** | **Injection site erythema** | **33272 (4.42%)** |
| TD ADSORBED (NO BRAND NAME) | 15.44% | DTAP (INFANRIX) | 13.54% |
| DTP (NO BRAND NAME) | 10.22% | PNEUMO (PNEUMOVAX) | 10.06% |
| DTAP (TRIPEDIA) | 7.02% | DTAP (TRIPEDIA) | 9.00% |
| **Rash** | **12034 (3.36%)** | **Erythema** | **20638 (2.74%)** |
| MEASLES + MUMPS + RUBELLA (MMR) | 14.21% | DTAP (INFANRIX) | 10.01% |
| VARICELLA (VARIVAX) | 10.55% | PNEUMO (PNEUMOVAX) | 9.68% |
| HEP B (ENGERIX-B) | 10.52% | DTAP (DAPTACEL) | 8.71% |
| **Injection site oedema** | **11268 (3.14%)** | **Injection site swelling** | **19202 (2.55%)** |
| TD ADSORBED (NO BRAND NAME) | 16.02% | PNEUMO (PNEUMOVAX) | 11.68% |
| DTP (NO BRAND NAME) | 11.73% | DTAP (INFANRIX) | 11.11% |
| DTAP (TRIPEDIA) | 7.26% | DTAP (DAPTACEL) | 8.04% |
| **Vasodilatation** | **10339 (2.88%)** | **Injection site pain** | **17325 (2.3%)** |
| TD ADSORBED (NO BRAND NAME) | 14.62% | PNEUMO (PNEUMOVAX) | 14.46% |
| DTP (NO BRAND NAME) | 8.78% | INFLUENZA (SEASONAL) (FLUZONE) | 10.05% |
| DTAP (TRIPEDIA) | 6.79% | HPV (GARDASIL) | 6.49% |
| 2010-2020 top 5 adverse events |  | 2021 top 5 adverse events |  |
| **Pyrexia** | **45536 (3.33%)** | **Headache** | **95058 (4.84%)** |
| ZOSTER (SHINGRIX) | 20.76% | COVID19 (COVID19 (MODERNA)) | 43.75% |
| PNEUMO (PNEUMOVAX) | 5.23% | COVID19 (COVID19 (PFIZER-BIONTECH)) | 40.77% |
| INFLUENZA (SEASONAL) (FLUZONE H | 3.97% | COVID19 (COVID19 (JANSSEN)) | 12.98% |
| **Injection site pain** | **38934 (2.85%)** | **Pyrexia** | **82643 (4.21%)** |
| ZOSTER (SHINGRIX) | 18.99% | COVID19 (COVID19 (MODERNA)) | 46.77% |
| PNEUMO (PNEUMOVAX) | 10.58% | COVID19 (COVID19 (PFIZER-BIONTECH)) | 37.11% |
| ZOSTER LIVE (ZOSTAVAX) | 5.67% | COVID19 (COVID19 (JANSSEN)) | 12.79% |
| **Injection site erythema** | **37276 (2.73%)** | **Fatigue** | **73491 (3.74%)** |
| ZOSTER LIVE (ZOSTAVAX) | 14.31% | COVID19 (COVID19 (MODERNA)) | 45.07% |
| PNEUMO (PNEUMOVAX) | 10.26% | COVID19 (COVID19 (PFIZER-BIONTECH)) | 41.77% |
| ZOSTER (SHINGRIX) | 6.22% | COVID19 (COVID19 (JANSSEN)) | 10.76% |
| **Incorrect product storage** | **29322 (2.15%)** | **Pain** | **71897 (3.66%)** |
| VARICELLA (VARIVAX) | 16.57% | COVID19 (COVID19 (MODERNA)) | 43.86% |
| ROTAVIRUS (ROTATEQ) | 12.27% | COVID19 (COVID19 (PFIZER-BIONTECH)) | 39.97% |
| MEASLES + MUMPS + RUBELLA + VAR | 8.79% | COVID19 (COVID19 (JANSSEN)) | 12.17% |
| **Injection site swelling** | **27930 (2.04%)** | **Nausea** | **55723 (2.84%)** |
| ZOSTER LIVE (ZOSTAVAX) | 12.09% | COVID19 (COVID19 (MODERNA)) | 43.68% |
| PNEUMO (PNEUMOVAX) | 12.00% | COVID19 (COVID19 (PFIZER-BIONTECH)) | 41.94% |
| ZOSTER (SHINGRIX) | 5.66% | COVID19 (COVID19 (JANSSEN)) | 11.75% |

### B. Nondomestic data

There were 346 210 nondomestic reports from 1990 through 2021. Reports included 214 vaccines. 2021 had the most reports (n = 243 500, 70.33%), followed by 2019 (n = 9,569, 2.76%), 2018 (n = 8,673, 2.512%), 2020 (n = 8,111, 2.34%), and 2016 (n = 7,991, 2.31%). Most of the sample (61.15%) came from female subjects, and the average age was 20.94 years. Supplementary figure 12 presents the number of reports per year. In addition, table 2 presents number of domestic and nondomestic reports per vaccine.

The top vaccine in terms of nondomestic reports is Pfizer-Biontech (57.64%), followed by Moderna (9.04%), Gardasil (2.57%), Janssen (2.48%), and seasonal influenza, no brand name (1.76%).

### Reported deaths

The total number of reported deaths is 14 125 (4.08%). The most deaths occurred in 1991 (10.17%), followed by 1993 (9.32%), 1994 (8.70%), 1990 (8.16%), and 1992 (8.00%). Supplementary figure 13 presents pattern of nondomestic reported death over years. The top vaccine in terms of reported deaths is Pfizer-Biontech (61.27%), followed by Moderna (6.77%), Janssen (3.88%), seasonal influenza, no brand name (2.43%), and Pneumovax (1.67%). Supplementary figure 14 shows the top vaccines in terms of reported deaths per year. In addition, supplementary table 10 presents the number of nondomestic reported deaths per vaccine per year.

### Reported ER or doctor office visits

The total number of reported ER or doctor office visits is 7,868 (2.27%), with the most ER or doctor visits occurring in 2015 (24.50%), followed by 2014 (19.83%), 2013 (18.07%), 2016 (17.72%), and 2000 (11.42%). Supplementary figure 15 shows pattern of nondomestic reported ER or doctor visits over years.

The top vaccine in terms of reported ER or doctor visits is Gardasil (21.59%), followed by Pneumovax (9.13%), Infanrix Hexa (5.31%), seasonal influenza, no brand name (4.49%), and Rotateq (4.31%). Supplementary figure 16 presents the pattern of top vaccines in reported ER or doctor visits over the years. In addition, supplementary table 11 presents the number of nondomestic reported ER or doctor visits per vaccine per year.

### Reported hospitalization

The total number of reported hospitalizations is 96 166 (27.78%) with hospitalization occurring most frequently in 1994 (76.40%), followed by 1993 (72.67%), 1997 (68.44%), 1995 (66.54%), and 1996 (65.07%). Supplementary figure 17 shows pattern of nondomestic reported hospitalization over the years.

The average number of hospitalization days in nondomestic reports was 6.10, with the longest average hospital stay occurring in 1992 (mean = 19.67).

Pfizer-Biontech is the top vaccine in terms of reported hospitalizations (45.80%), followed by Moderna (8.12%), Gardasil (3.36%), Pneumovax (2.46%), and Infanrix Hexa (2.32%). Supplementary figure 18 presents patterns of the top vaccines in terms of nondomestic reported hospitalization. In addition, supplementary table 12 presents the number of nondomestic reported hospitalizations per vaccine per year.

### Reported lethal threats

The number of reported lethal threats is 15 151 (4.38%), with the most lethal threats occurring in 1992 (17.60%), followed by 1991 (10.17%), 1994 (9.94%), 1995 (6.99%), and 2000 (6.85%). Supplementary figure 19 presents the pattern of nondomestic reported lethal threat over the years.

COVID-19 vaccines were the top vaccines in terms of reported lethal threats—Pfizer-Biontech (60.89%), Moderna (9.16%), and Janssen (2.83%)—followed by Infanrix Hexa (1.89%) and seasonal influenza, no brand name (1.80%). Supplementary figure 20 presents the top vaccines in terms of nondomestic lethal threats. In addition, supplementary table 13 presents the number of nondomestic reported lethal threats per vaccine per year.

### Reported disabilities

The total number of reported disabilities is 27 545 (7.96%), with reported disabilities occurring most frequently in 1990 (22.45%), followed by 2001 (18.21%), 2002 (16.71%), 1999 (15.76%), and 2003 (14.59%). Supplementary figure 21 presents pattern of nondomestic reported disabilities over the years.

Pfizer-Biontech is the top vaccine in terms of nondomestic reported disabilities (59.52%), followed by Moderna (10.36%), Gardasil (4.76%), Engerix-B (3.42%), and Janssen (2.70%). Figure 22 presents the top vaccines in terms of nondomestic reported disabilities over the years. In addition, supplementary table 14 presents the number of nondomestic reported hospitalizations per vaccine per year.

### Most frequently reported nondomestic reported symptoms over the years

The total number of nondomestic reported symptoms is 2 159 135. Table 15 presents the top vaccines in terms of reported symptoms. From 1990 through 2000, pyrexia was the most frequently reported AEFI (5.27%), followed by infection (2.44%), abnormal laboratory test (2.4%), convulsion (1.88%), and vomiting (1.49%). Pyrexia was also the most frequently reported AEFI (4.28%) from 2001 through 2010, followed by vomiting (1.51%), increased body temperature (1.47%), crying (1.41%), and pallor (1.37%). From 2011 through 2021, pyrexia was again the most frequently reported AEFI (3.8%), followed by vomiting (1.40%), headache (1.18%), crying (1.16%), and diarrhea (0.96%). Finally, in 2021, the most frequently reported AEFI was SARS-CoV-2 test (4.59%), followed by headache (3.18%), fatigue (2.57%), pyrexia (2.55%), and nausea (1.82%). Table 16 presents the most frequently reported AEFIs with the most frequently associated vaccines.

**Table 15:** Top vaccines in non-domestic reported symptoms

| **Vaccines** | **%** | **Count of reported symptoms** |
| --- | --- | --- |
| **COVID19 (COVID19 (PFIZER-BIONTECH))** | 50.84% | 1097734 |
| **COVID19 (COVID19 (MODERNA))** | 6.34% | 136895 |
| **HPV (GARDASIL)** | 3.21% | 69362 |
| **PNEUMO (PREVNAR13)** | 2.82% | 60813 |
| **ROTAVIRUS (ROTARIX)** | 1.80% | 38758 |
| **INFLUENZA (SEASONAL) (NO BRAND NAME)** | 1.79% | 38702 |
| **HEP B (ENGRIX-B)** | 1.78% | 38499 |
| **PNEUMO (PNEUMOVAX)** | 1.51% | 32520 |
| **COVID19 (COVID19 (JANSSEN))** | 1.48% | 32017 |
| **HPV (CERVARIX)** | 1.39% | 30053 |
| **PNEUMO (PREVNAR)** | 1.38% | 29748 |
| **HIB (ACTHIB)** | 1.25% | 26980 |
| **DTAP+IPV+HEPB+HIB (INFANRIX HEXA)** | 1.22% | 26376 |
| **MEASLES + MUMPS + RUBELLA (MMR II)** | 1.13% | 24327 |
| **HEP B (NO BRAND NAME)** | 0.99% | 21418 |
| **POLIO VIRUS, INACT. (NO BRAND NAME)** | 0.94% | 20402 |
| **ROTAVIRUS (ROTATEQ)** | 0.92% | 19841 |
| **DTAP (INFANRIX)** | 0.90% | 19480 |
| **MENINGOCOCCAL B (BEXSERO)** | 0.89% | 19113 |
| **MEASLES + MUMPS + RUBELLA (NO BRAND NAME)** | 0.82% | 17721 |
| **Total** | 100% | 2159117 |

**Table 16:** Top non-domestic reported AEFI with top related vaccines

| 1990-2000 top 5 adverse events |  | 2001-2010 top 5 adverse events |  |
| --- | --- | --- | --- |
| **Pyrexia** | **1439 (5.27%)** | **Pyrexia** | **9241 (4.28%)** |
| MEASLES + MUMPS + RUBELLA (MMR II) | 14.18% | PNEUMO (PREVNAR) | 10.89% |
| HEP B (ENGERIX-B) | 12.37% | POLIO VIRUS, INACT. (NO BRAND NAME) | 7.94% |
| DTP (NO BRAND NAME) | 6.67% | DTAP (INFANRIX) | 7.72% |
| **Infection** | **1439 (2.44%)** | **Vomiting** | **3260 (1.51%)** |
| HEP B (ENGERIX-B) | 20.21% | PNEUMO (PREVNAR) | 9.54% |
| MEASLES + MUMPS + RUBELLA (MMR II) | 10.03% | ROTAVIRUS (ROTARIX) | 9.26% |
| HEP B (RECOMBIVAX HB) | 7.49% | POLIO VIRUS, INACT. (NO BRAND NAME) | 6.69% |
| **Laboratory test abnormal** | **655 (2.4%)** | **Body temperature increased** | **3185 (1.47%)** |
| HEP B (ENGERIX-B) | 38.02% | PNEUMO (PREVNAR) | 16.99% |
| HEP B (RECOMBIVAX HB) | 9.16% | POLIO VIRUS, INACT. (NO BRAND NAME) | 9.92% |
| MEASLES + MUMPS + RUBELLA (MMR II) | 8.70% | DTAP (INFANRIX) | 8.98% |
| **Convulsion** | **513 (1.88%)** | **Crying** | **3051 (1.41%)** |
| MEASLES + MUMPS + RUBELLA (MMR II) | 11.89% | PNEUMO (PREVNAR) | 13.67% |
| HEP B (ENGERIX-B) | 11.11% | POLIO VIRUS, INACT. (NO BRAND NAME) | 10.62% |
| HEP B (RECOMBIVAX HB) | 8.19% | HIB (ACTHIB) | 9.73% |
| **Vomiting** | **407 (1.49%)** | **Pallor** | **2958 (1.37%)** |
| HEP B (ENGERIX-B) | 17.44% | PNEUMO (PREVNAR) | 13.56% |
| MEASLES + MUMPS + RUBELLA (MMR II) | 10.32% | POLIO VIRUS, INACT. (NO BRAND NAME) | 12.04% |
| DTP (NO BRAND NAME) | 6.88% | DTAP (INFANRIX) | 10.92% |
| 2011-2020 top 5 adverse events |  | 2021 top 5 adverse events |  |
| **Pyrexia** | **23397 (3.8%)** | **SARS-CoV-2 test** | **59666 (4.59%)** |
| PNEUMO (PREVNAR13) | 10.83% | COVID19 (PFIZER-BIONTECH) | 88.86% |
| MENINGOCOCCAL B (BEXSERO) | 5.88% | COVID19 (MODERNA) | 9.24% |
| PNEUMO (PNEUMOVAX) | 5.58% | COVID19 (JANSSEN) | 1.50% |
| **Vomiting** | **8650 (1.40%)** | **Headache** | **41312 (3.18%)** |
| ROTAVIRUS (ROTARIX) | 13.03% | COVID19 (PFIZER-BIONTECH) | 80.10% |
| PNEUMO (PREVNAR13) | 9.23% | COVID19 (MODERNA) | 14.96% |
| ROTAVIRUS (ROTATEQ) | 6.10% | COVID19 (JANSSEN) | 3.83% |
| **Headache** | **7266 (1.18%)** | **Fatigue** | **33405 (2.57%)** |
| HPV (GARDASIL) | 27.00% | COVID19 (PFIZER-BIONTECH) | 81.10% |
| HPV (CERVARIX) | 10.38% | COVID19 (MODERNA) | 14.64% |
| INFLUENZA (SEASONAL) (NO BRAND NAME) | 5.78% | COVID19 (JANSSEN) | 3.23% |
| **Crying** | **7149 (1.16%)** | **Pyrexia** | **33118 (2.55%)** |
| PNEUMO (PREVNAR13) | 13.50% | COVID19 (PFIZER-BIONTECH) | 71.93% |
| ROTAVIRUS (ROTARIX) | 9.22% | COVID19 (MODERNA) | 20.01% |
| HIB (ACTHIB) | 8.71% | COVID19 (JANSSEN) | 4.19% |
| **Diarrhoea** | **5913 (0.96%)** | **Nausea** | **23607 (1.82%)** |
| ROTAVIRUS (ROTARIX) | 18.03% | COVID19 (PFIZER-BIONTECH) | 79.76% |
| PNEUMO (PREVNAR13) | 9.25% | COVID19 (MODERNA) | 15.73% |
| ROTAVIRUS (ROTATEQ) | 8.30% | COVID19 (JANSSEN) | 3.34% |

### C. Survival Analysis Results

Domestic reports with complete vaccination and onset data are 1 121 698, while for nondomestic data they are 262 723. Probability of having non serious AEFIs has its highest value at the vaccination’s day, with 93% probability of having non serious AEFIs in domestic data, and 85.91% in nondomestic data. It continuously decreased after that to its minimal 10.40% and 6.08% at the end of the following up window, for domestic and nondomestic data, respectively. However, for serious AEFIs they have a proportional relation to time, where they have their lowest values in the vaccination’s day, ranging from 0.09% to 5.18% for domestic data and ranging from 0.39% to 8.00% for nondomestic data. For domestic data ER or Doctor visit was the highest serious AEFIs by 33.84%, and for nondomestic data hospitalization was the highest by 54.10% at the end of the follow up window. Table 17 contains survival analysis of both domestic and nondomestic data. In addition, supplementary table 18 contains cox regression results of both domestic and nondomestic data. Both in domestic and nondomestic data, males have more probability to have AEFIs than females (HR: 1.15, 1.14 - 1.16) in domestic, and (HR: 1.23, 1.22 -1.25) in nondomestic. While for age group, in domestic data age group ≤ 5 years have a higher probability of having AEFIs compared to other groups. However, in nondomestic data age group > 84 years have a higher probability of having AEFIs compared to other groups (HR:2. 15, 2.07 - 2.23).

**Table 17:** Survival analysis of domestic and nondomestic data

| Domestic Data Survival Analysis |  |  |  |  |  |  |  |  |  |  |  |  |
| --- | --- | --- | --- | --- | --- | --- | --- | --- | --- | --- | --- | --- |
| time(days) | n.risk | n.event | Survival<br>or<br>P(non serious<br>AEFIs) | std.err | lower<br>95% CI | upper<br>95% CI | P (Death) | P (ER or<br>Doctor<br>visit) | P (Hospitalization) | P (Lethal<br>threat) | P (Disability) | P (Multiple<br>serious AEFIs) |
| 0 | 1112242 | 78028 | 0.93 | 0.000242 | 0.929 | 0.93 | 0.000899 | 0.0518 | 0.00593 | 0.00121 | 0.0024 | 0.00795 |
| 1 | 597977 | 49420 | 0.853 | 0.000399 | 0.852 | 0.854 | 0.00316 | 0.1075 | 0.01368 | 0.00186 | 0.00475 | 0.01604 |
| 2 | 354436 | 19049 | 0.807 | 0.000497 | 0.806 | 0.808 | 0.004732 | 0.1384 | 0.0194 | 0.00224 | 0.00636 | 0.02173 |
| 3 | 281684 | 9043 | 0.781 | 0.00055 | 0.78 | 0.782 | 0.005918 | 0.1524 | 0.02405 | 0.00254 | 0.00759 | 0.02626 |
| 4 | 248551 | 5601 | 0.764 | 0.000586 | 0.762 | 0.765 | 0.006833 | 0.1611 | 0.02723 | 0.00275 | 0.00849 | 0.03 |
| 5 | 228377 | 4565 | 0.748 | 0.000616 | 0.747 | 0.75 | 0.007608 | 0.1686 | 0.03008 | 0.00291 | 0.00922 | 0.03319 |
| 6 | 212285 | 3840 | 0.735 | 0.000643 | 0.734 | 0.736 | 0.008296 | 0.1756 | 0.03241 | 0.00312 | 0.00978 | 0.03596 |
| 7 | 197700 | 5425 | 0.715 | 0.000681 | 0.713 | 0.716 | 0.009076 | 0.1865 | 0.03561 | 0.00335 | 0.0109 | 0.03994 |
| 30 | 60908 | 33564 | 0.521 | 0.001061 | 0.519 | 0.523 | 0.019265 | 0.2612 | 0.08247 | 0.00624 | 0.02228 | 0.08751 |
| 60 | 37745 | 7723 | 0.442 | 0.001227 | 0.44 | 0.444 | 0.02439 | 0.2783 | 0.10768 | 0.00762 | 0.02846 | 0.11146 |
| 90 | 29066 | 3299 | 0.4 | 0.001311 | 0.397 | 0.403 | 0.026516 | 0.288 | 0.12232 | 0.00866 | 0.03126 | 0.12327 |
| 100 | 26608 | 895 | 0.387 | 0.001337 | 0.385 | 0.39 | 0.027101 | 0.2908 | 0.1273 | 0.00888 | 0.03201 | 0.12662 |
| 200 | 8884 | 8045 | 0.232 | 0.001607 | 0.229 | 0.235 | 0.035029 | 0.3098 | 0.21957 | 0.0103 | 0.03639 | 0.15691 |
| 300 | 1292 | 3378 | 0.104 | 0.001816 | 0.101 | 0.108 | 0.041138 | 0.3384 | 0.292 | 0.01099 | 0.03944 | 0.17401 |
| Non-Domestic Data Survival Analysis |  |  |  |  |  |  |  |  |  |  |  |  |
| time(days) | n.risk | n.event | Survival<br>or<br>P(non serious<br>AEFIs) | std.err | lower<br>95% CI | upper<br>95% CI | P (Death) | P (ER or<br>Doctor<br>visit) | P (Hospitalization) | P (Lethal<br>threat) | P (Disability) | P (Multiple<br>serious AEFIs) |
| 0 | 260378 | 36695 | 0.8591 | 0.000682 | 0.8577 | 0.8604 | 0.00577 | 0.00394 | 0.08 | 0.00893 | 0.0262 | 0.016 |
| 1 | 155488 | 16485 | 0.768 | 0.000906 | 0.7662 | 0.7698 | 0.01325 | 0.00566 | 0.127 | 0.01224 | 0.0478 | 0.0262 |
| 2 | 112050 | 7030 | 0.7198 | 0.001015 | 0.7178 | 0.7218 | 0.0174 | 0.00659 | 0.154 | 0.01424 | 0.0551 | 0.0325 |
| 3 | 96014 | 4934 | 0.6828 | 0.001091 | 0.6807 | 0.685 | 0.02071 | 0.00716 | 0.176 | 0.01552 | 0.0602 | 0.0379 |
| 4 | 85967 | 3416 | 0.6557 | 0.001142 | 0.6534 | 0.6579 | 0.02278 | 0.00758 | 0.192 | 0.01654 | 0.0635 | 0.0422 |
| 5 | 79038 | 2818 | 0.6323 | 0.001184 | 0.63 | 0.6346 | 0.02492 | 0.00789 | 0.205 | 0.01735 | 0.0661 | 0.046 |
| 6 | 73505 | 2319 | 0.6124 | 0.001217 | 0.61 | 0.6147 | 0.02648 | 0.00821 | 0.218 | 0.01799 | 0.0685 | 0.049 |
| 7 | 68875 | 2691 | 0.5884 | 0.001253 | 0.586 | 0.5909 | 0.02814 | 0.00854 | 0.232 | 0.0189 | 0.0716 | 0.0525 |
| 30 | 26492 | 19339 | 0.3714 | 0.001497 | 0.3685 | 0.3744 | 0.04357 | 0.01082 | 0.363 | 0.02763 | 0.0947 | 0.0893 |
| 60 | 13739 | 5328 | 0.2819 | 0.001569 | 0.2789 | 0.285 | 0.04853 | 0.01203 | 0.415 | 0.03131 | 0.1046 | 0.106 |
| 90 | 8299 | 1996 | 0.2339 | 0.001635 | 0.2307 | 0.2371 | 0.05143 | 0.01271 | 0.443 | 0.03265 | 0.1103 | 0.1157 |
| 100 | 7181 | 419 | 0.2215 | 0.001657 | 0.2183 | 0.2248 | 0.05217 | 0.01301 | 0.45 | 0.03295 | 0.1118 | 0.1184 |
| 200 | 1925 | 2058 | 0.133 | 0.00189 | 0.1294 | 0.1368 | 0.05906 | 0.01439 | 0.502 | 0.03539 | 0.1185 | 0.1375 |
| 300 | 417 | 719 | 0.0608 | 0.00213 | 0.0567 | 0.0651 | 0.06389 | 0.01662 | 0.541 | 0.03675 | 0.1275 | 0.1538 |

### D. Adverse event of special interest (AESIs)

Out of 3 449 887 domestic reported symptoms 17 611 (0.51%) were AESIs, 9 665 (54.88%) were associated with COVID-19 vaccines. Table 19 shows distribution of domestic AESIs associated with COVID-19 stratified by gender and age. While for nondomestic reported symptoms out of 2 159 135 there were 35 810 (1.66%) AESIs, 31 511 (87.99%) were associated with COVID-19 vaccines. Table 20 shows distribution of nondomestic AESIs associated with COVID-19 stratified by gender and age.

**Table 19:**
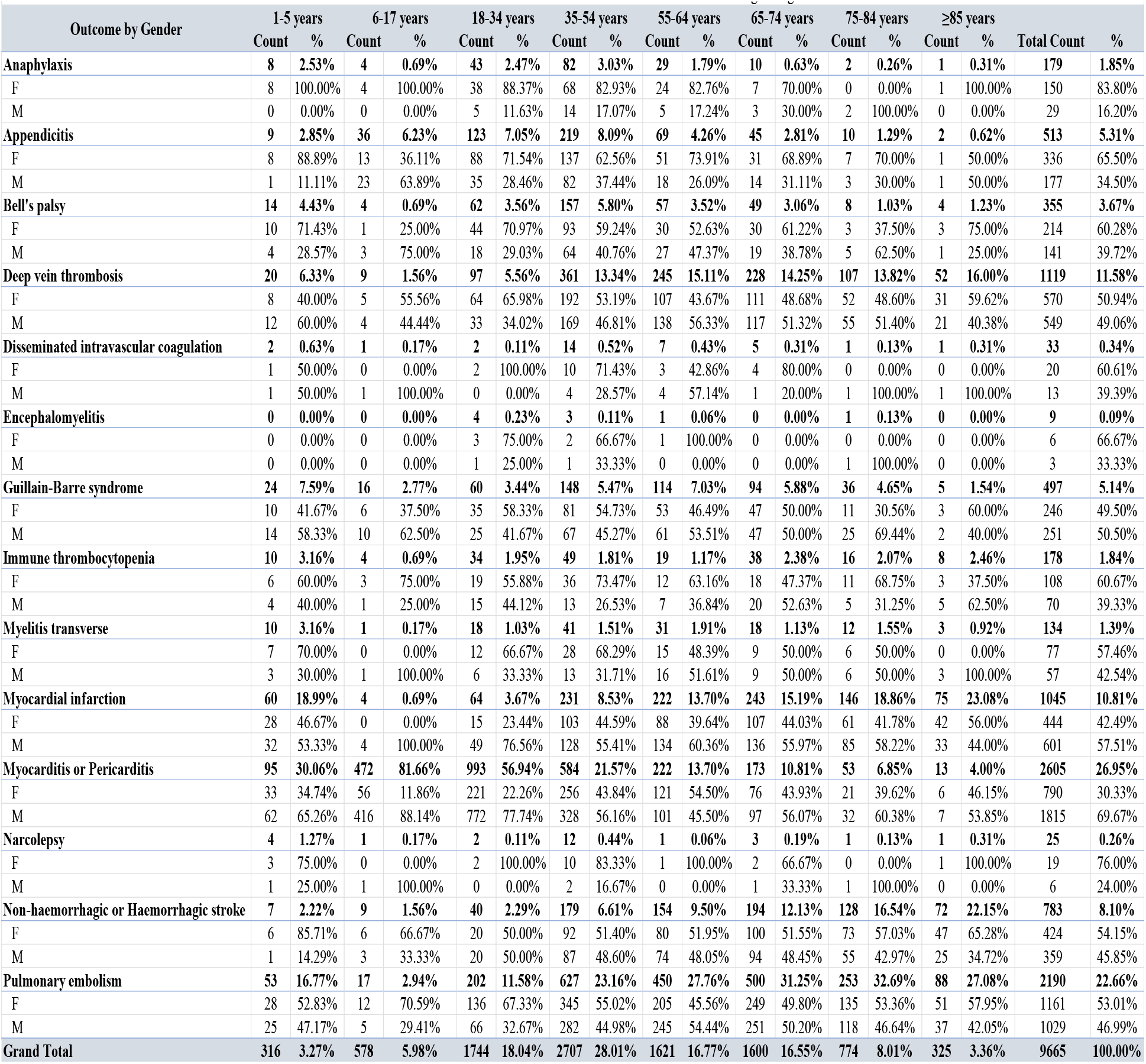
Survival analysis of domestic and nondomestic data

| Outcome by Gender | 1-5 years |  | 6-17 years |  | 18-34 years |  | 35-54 years |  | 55-64 years |  | 65-74 years |  | 75-84 years |  | ≥85 years |  | Total Count | % |
| --- | --- | --- | --- | --- | --- | --- | --- | --- | --- | --- | --- | --- | --- | --- | --- | --- | --- | --- |
|  | Count | % | Count | % | Count | % | Count | % | Count | % | Count | % | Count | % | Count | % |  |  |
| **Anaphylaxis** | **8** | **2.53%** | **4** | **0.69%** | **43** | **2.47%** | **82** | **3.03%** | **29** | **1.79%** | **10** | **0.63%** | **2** | **0.26%** | **1** | **0.31%** | **179** | **1.85%** |
| F | 8 | 100.00% | 4 | 100.00% | 38 | 88.37% | 68 | 82.93% | 24 | 82.76% | 7 | 70.00% | 0 | 0.00% | 1 | 100.00% | 150 | 83.80% |
| M | 0 | 0.00% | 0 | 0.00% | 5 | 11.63% | 14 | 17.07% | 5 | 17.24% | 3 | 30.00% | 2 | 100.00% | 0 | 0.00% | 29 | 16.20% |
| **Appendicitis** | **9** | **2.85%** | **36** | **6.23%** | **123** | **7.05%** | **219** | **8.09%** | **69** | **4.26%** | **45** | **2.81%** | **10** | **1.29%** | **2** | **0.62%** | **513** | **5.31%** |
| F | 8 | 88.89% | 13 | 36.11% | 88 | 71.54% | 137 | 62.56% | 51 | 73.91% | 31 | 68.89% | 7 | 70.00% | 1 | 50.00% | 336 | 65.50% |
| M | 1 | 11.11% | 23 | 63.89% | 35 | 28.46% | 82 | 37.44% | 18 | 26.09% | 14 | 31.11% | 3 | 30.00% | 1 | 50.00% | 177 | 34.50% |
| **Bell's palsy** | **14** | **4.43%** | **4** | **0.69%** | **62** | **3.56%** | **157** | **5.80%** | **57** | **3.52%** | **49** | **3.06%** | **8** | **1.03%** | **4** | **1.23%** | **355** | **3.67%** |
| F | 10 | 71.43% | 1 | 25.00% | 44 | 70.97% | 93 | 59.24% | 30 | 52.63% | 30 | 61.22% | 3 | 37.50% | 3 | 75.00% | 214 | 60.28% |
| M | 4 | 28.57% | 3 | 75.00% | 18 | 29.03% | 64 | 40.76% | 27 | 47.37% | 19 | 38.78% | 5 | 62.50% | 1 | 25.00% | 141 | 39.72% |
| **Deep vein thrombosis** | **20** | **6.33%** | **9** | **1.56%** | **97** | **5.56%** | **361** | **13.34%** | **245** | **15.11%** | **228** | **14.25%** | **107** | **13.82%** | **52** | **16.00%** | **1119** | **11.58%** |
| F | 8 | 40.00% | 5 | 55.56% | 64 | 65.98% | 192 | 53.19% | 107 | 43.67% | 111 | 48.68% | 52 | 48.60% | 31 | 59.62% | 570 | 50.94% |
| M | 12 | 60.00% | 4 | 44.44% | 33 | 34.02% | 169 | 46.81% | 138 | 56.33% | 117 | 51.32% | 55 | 51.40% | 21 | 40.38% | 549 | 49.06% |
| **Disseminated intravascular coagulation** | **2** | **0.63%** | **1** | **0.17%** | **2** | **0.11%** | **14** | **0.52%** | **7** | **0.43%** | **5** | **0.31%** | **1** | **0.13%** | **1** | **0.31%** | **33** | **0.34%** |
| F | 1 | 50.00% | 0 | 0.00% | 2 | 100.00% | 10 | 71.43% | 3 | 42.86% | 4 | 80.00% | 0 | 0.00% | 0 | 0.00% | 20 | 60.61% |
| M | 1 | 50.00% | 1 | 100.00% | 0 | 0.00% | 4 | 28.57% | 4 | 57.14% | 1 | 20.00% | 1 | 100.00% | 1 | 100.00% | 13 | 39.39% |
| **Encephalomyelitis** | **0** | **0.00%** | **0** | **0.00%** | **4** | **0.23%** | **3** | **0.11%** | **1** | **0.06%** | **0** | **0.00%** | **1** | **0.13%** | **0** | **0.00%** | **9** | **0.09%** |
| F | 0 | 0.00% | 0 | 0.00% | 3 | 75.00% | 2 | 66.67% | 1 | 100.00% | 0 | 0.00% | 0 | 0.00% | 0 | 0.00% | 6 | 66.67% |
| M | 0 | 0.00% | 0 | 0.00% | 1 | 25.00% | 1 | 33.33% | 0 | 0.00% | 0 | 0.00% | 1 | 100.00% | 0 | 0.00% | 3 | 33.33% |
| **Guillain-Barre syndrome** | **24** | **7.59%** | **16** | **2.77%** | **60** | **3.44%** | **148** | **5.47%** | **114** | **7.03%** | **94** | **5.88%** | **36** | **4.65%** | **5** | **1.54%** | **497** | **5.14%** |
| F | 10 | 41.67% | 6 | 37.50% | 35 | 58.33% | 81 | 54.73% | 53 | 46.49% | 47 | 50.00% | 11 | 30.56% | 3 | 60.00% | 246 | 49.50% |
| M | 14 | 58.33% | 10 | 62.50% | 25 | 41.67% | 67 | 45.27% | 61 | 53.51% | 47 | 50.00% | 25 | 69.44% | 2 | 40.00% | 251 | 50.50% |
| **Immune thrombocytopenia** | **10** | **3.16%** | **4** | **0.69%** | **34** | **1.95%** | **49** | **1.81%** | **19** | **1.17%** | **38** | **2.38%** | **16** | **2.07%** | **8** | **2.46%** | **178** | **1.84%** |
| F | 6 | 60.00% | 3 | 75.00% | 19 | 55.88% | 36 | 73.47% | 12 | 63.16% | 18 | 47.37% | 11 | 68.75% | 3 | 37.50% | 108 | 60.67% |
| M | 4 | 40.00% | 1 | 25.00% | 15 | 44.12% | 13 | 26.53% | 7 | 36.84% | 20 | 52.63% | 5 | 31.25% | 5 | 62.50% | 70 | 39.33% |
| **Myelitis transverse** | **10** | **3.16%** | **1** | **0.17%** | **18** | **1.03%** | **41** | **1.51%** | **31** | **1.91%** | **18** | **1.13%** | **12** | **1.55%** | **3** | **0.92%** | **134** | **1.39%** |
| F | 7 | 70.00% | 0 | 0.00% | 12 | 66.67% | 28 | 68.29% | 15 | 48.39% | 9 | 50.00% | 6 | 50.00% | 0 | 0.00% | 77 | 57.46% |
| M | 3 | 30.00% | 1 | 100.00% | 6 | 33.33% | 13 | 31.71% | 16 | 51.61% | 9 | 50.00% | 6 | 50.00% | 3 | 100.00% | 57 | 42.54% |
| **Myocardial infarction** | **60** | **18.99%** | **4** | **0.69%** | **64** | **3.67%** | **231** | **8.53%** | **222** | **13.70%** | **243** | **15.19%** | **146** | **18.86%** | **75** | **23.08%** | **1045** | **10.81%** |
| F | 28 | 46.67% | 0 | 0.00% | 15 | 23.44% | 103 | 44.59% | 88 | 39.64% | 107 | 44.03% | 61 | 41.78% | 42 | 56.00% | 444 | 42.49% |
| M | 32 | 53.33% | 4 | 100.00% | 49 | 76.56% | 128 | 55.41% | 134 | 60.36% | 136 | 55.97% | 85 | 58.22% | 33 | 44.00% | 601 | 57.51% |
| **Myocarditis or Pericarditis** | **95** | **30.06%** | **472** | **81.66%** | **993** | **56.94%** | **584** | **21.57%** | **222** | **13.70%** | **173** | **10.81%** | **53** | **6.85%** | **13** | **4.00%** | **2605** | **26.95%** |
| F | 33 | 34.74% | 56 | 11.86% | 221 | 22.26% | 256 | 43.84% | 121 | 54.50% | 76 | 43.93% | 21 | 39.62% | 6 | 46.15% | 790 | 30.33% |
| M | 62 | 65.26% | 416 | 88.14% | 772 | 77.74% | 328 | 56.16% | 101 | 45.50% | 97 | 56.07% | 32 | 60.38% | 7 | 53.85% | 1815 | 69.67% |
| **Narcolepsy** | **4** | **1.27%** | **1** | **0.17%** | **2** | **0.11%** | **12** | **0.44%** | **1** | **0.06%** | **3** | **0.19%** | **1** | **0.13%** | **1** | **0.31%** | **25** | **0.26%** |
| F | 3 | 75.00% | 0 | 0.00% | 2 | 100.00% | 10 | 83.33% | 1 | 100.00% | 2 | 66.67% | 0 | 0.00% | 1 | 100.00% | 19 | 76.00% |
| M | 1 | 25.00% | 1 | 100.00% | 0 | 0.00% | 2 | 16.67% | 0 | 0.00% | 1 | 33.33% | 1 | 100.00% | 0 | 0.00% | 6 | 24.00% |
| **Non-haemorrhagic or Haemorrhagic stroke** | **7** | **2.22%** | **9** | **1.56%** | **40** | **2.29%** | **179** | **6.61%** | **154** | **9.50%** | **194** | **12.13%** | **128** | **16.54%** | **72** | **22.15%** | **783** | **8.10%** |
| F | 6 | 85.71% | 6 | 66.67% | 20 | 50.00% | 92 | 51.40% | 80 | 51.95% | 100 | 51.55% | 73 | 57.03% | 47 | 65.28% | 424 | 54.15% |
| M | 1 | 14.29% | 3 | 33.33% | 20 | 50.00% | 87 | 48.60% | 74 | 48.05% | 94 | 48.45% | 55 | 42.97% | 25 | 34.72% | 359 | 45.85% |
| **Pulmonary embolism** | **53** | **16.77%** | **17** | **2.94%** | **202** | **11.58%** | **627** | **23.16%** | **450** | **27.76%** | **500** | **31.25%** | **253** | **32.69%** | **88** | **27.08%** | **2190** | **22.66%** |
| F | 28 | 52.83% | 12 | 70.59% | 136 | 67.33% | 345 | 55.02% | 205 | 45.56% | 249 | 49.80% | 135 | 53.36% | 51 | 57.95% | 1161 | 53.01% |
| M | 25 | 47.17% | 5 | 29.41% | 66 | 32.67% | 282 | 44.98% | 245 | 54.44% | 251 | 50.20% | 118 | 46.64% | 37 | 42.05% | 1029 | 46.99% |
| **Grand Total** | **316** | **3.27%** | **578** | **5.98%** | **1744** | **18.04%** | **2707** | **28.01%** | **1621** | **16.77%** | **1600** | **16.55%** | **774** | **8.01%** | **325** | **3.36%** | **9665** | **100.00%** |

**Table 20:**
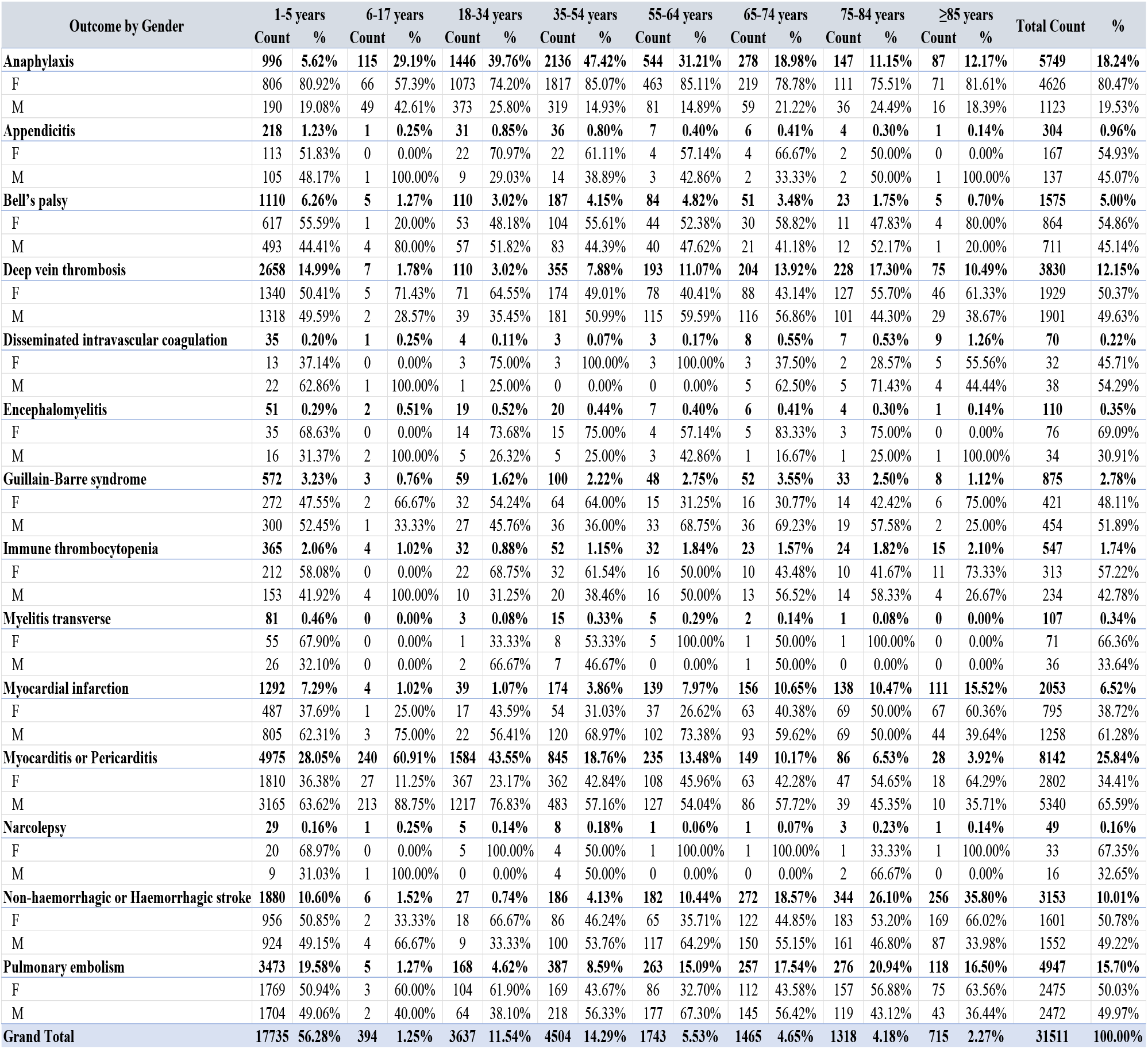
Survival analysis of domestic and nondomestic data

| Outcome by Gender | 1-5 years |  | 6-17 years |  | 18-34 years |  | 35-54 years |  | 55-64 years |  | 65-74 years |  | 75-84 years |  | ≥85 years |  | Total Count | % |
| --- | --- | --- | --- | --- | --- | --- | --- | --- | --- | --- | --- | --- | --- | --- | --- | --- | --- | --- |
|  | Count | % | Count | % | Count | % | Count | % | Count | % | Count | % | Count | % | Count | % |  |  |
| **Anaphylaxis** | **996** | **5.62%** | **115** | **29.19%** | **1446** | **39.76%** | **2136** | **47.42%** | **544** | **31.21%** | **278** | **18.98%** | **147** | **11.15%** | **87** | **12.17%** | **5749** | **18.24%** |
| F | 806 | 80.92% | 66 | 57.39% | 1073 | 74.20% | 1817 | 85.07% | 463 | 85.11% | 219 | 78.78% | 111 | 75.51% | 71 | 81.61% | 4626 | 80.47% |
| M | 190 | 19.08% | 49 | 42.61% | 373 | 25.80% | 319 | 14.93% | 81 | 14.89% | 59 | 21.22% | 36 | 24.49% | 16 | 18.39% | 1123 | 19.53% |
| **Appendicitis** | **218** | **1.23%** | **1** | **0.25%** | **31** | **0.85%** | **36** | **0.80%** | **7** | **0.40%** | **6** | **0.41%** | **4** | **0.30%** | **1** | **0.14%** | **304** | **0.96%** |
| F | 113 | 51.83% | 0 | 0.00% | 22 | 70.97% | 22 | 61.11% | 4 | 57.14% | 4 | 66.67% | 2 | 50.00% | 0 | 0.00% | 167 | 54.93% |
| M | 105 | 48.17% | 1 | 100.00% | 9 | 29.03% | 14 | 38.89% | 3 | 42.86% | 2 | 33.33% | 2 | 50.00% | 1 | 100.00% | 137 | 45.07% |
| **Bell's palsy** | **1110** | **6.26%** | **5** | **1.27%** | **110** | **3.02%** | **187** | **4.15%** | **84** | **4.82%** | **51** | **3.48%** | **23** | **1.75%** | **5** | **0.70%** | **1575** | **5.00%** |
| F | 617 | 55.59% | 1 | 20.00% | 53 | 48.18% | 104 | 55.61% | 44 | 52.38% | 30 | 58.82% | 11 | 47.83% | 4 | 80.00% | 864 | 54.86% |
| M | 493 | 44.41% | 4 | 80.00% | 57 | 51.82% | 83 | 44.39% | 40 | 47.62% | 21 | 41.18% | 12 | 52.17% | 1 | 20.00% | 711 | 45.14% |
| **Deep vein thrombosis** | **2658** | **14.99%** | **7** | **1.78%** | **110** | **3.02%** | **355** | **7.88%** | **193** | **11.07%** | **204** | **13.92%** | **228** | **17.30%** | **75** | **10.49%** | **3830** | **12.15%** |
| F | 1340 | 50.41% | 5 | 71.43% | 71 | 64.55% | 174 | 49.01% | 78 | 40.41% | 88 | 43.14% | 127 | 55.70% | 46 | 61.33% | 1929 | 50.37% |
| M | 1318 | 49.59% | 2 | 28.57% | 39 | 35.45% | 181 | 50.99% | 115 | 59.59% | 116 | 56.86% | 101 | 44.30% | 29 | 38.67% | 1901 | 49.63% |
| **Disseminated intravascular coagulation** | **35** | **0.20%** | **1** | **0.25%** | **4** | **0.11%** | **3** | **0.07%** | **3** | **0.17%** | **8** | **0.55%** | **7** | **0.53%** | **9** | **1.26%** | **70** | **0.22%** |
| F | 13 | 37.14% | 0 | 0.00% | 3 | 75.00% | 3 | 100.00% | 3 | 100.00% | 3 | 37.50% | 2 | 28.57% | 5 | 55.56% | 32 | 45.71% |
| M | 22 | 62.86% | 1 | 100.00% | 1 | 25.00% | 0 | 0.00% | 0 | 0.00% | 5 | 62.50% | 5 | 71.43% | 4 | 44.44% | 38 | 54.29% |
| **Encephalomyelitis** | **51** | **0.29%** | **2** | **0.51%** | **19** | **0.52%** | **20** | **0.44%** | **7** | **0.40%** | **6** | **0.41%** | **4** | **0.30%** | **1** | **0.14%** | **110** | **0.35%** |
| F | 35 | 68.63% | 0 | 0.00% | 14 | 73.68% | 15 | 75.00% | 4 | 57.14% | 5 | 83.33% | 3 | 75.00% | 0 | 0.00% | 76 | 69.09% |
| M | 16 | 31.37% | 2 | 100.00% | 5 | 26.32% | 5 | 25.00% | 3 | 42.86% | 1 | 16.67% | 1 | 25.00% | 1 | 100.00% | 34 | 30.91% |
| **Guillain-Barre syndrome** | **572** | **3.23%** | **3** | **0.76%** | **59** | **1.62%** | **100** | **2.22%** | **48** | **2.75%** | **52** | **3.55%** | **33** | **2.50%** | **8** | **1.12%** | **875** | **2.78%** |
| F | 272 | 47.55% | 2 | 66.67% | 32 | 54.24% | 64 | 64.00% | 15 | 31.25% | 16 | 30.77% | 14 | 42.42% | 6 | 75.00% | 421 | 48.11% |
| M | 300 | 52.45% | 1 | 33.33% | 27 | 45.76% | 36 | 36.00% | 33 | 68.75% | 36 | 69.23% | 19 | 57.58% | 2 | 25.00% | 454 | 51.89% |
| **Immune thrombocytopenia** | **365** | **2.06%** | **4** | **1.02%** | **32** | **0.88%** | **52** | **1.15%** | **32** | **1.84%** | **23** | **1.57%** | **24** | **1.82%** | **15** | **2.10%** | **547** | **1.74%** |
| F | 212 | 58.08% | 0 | 0.00% | 22 | 68.75% | 32 | 61.54% | 16 | 50.00% | 10 | 43.48% | 10 | 41.67% | 11 | 73.33% | 313 | 57.22% |
| M | 153 | 41.92% | 4 | 100.00% | 10 | 31.25% | 20 | 38.46% | 16 | 50.00% | 13 | 56.52% | 14 | 58.33% | 4 | 26.67% | 234 | 42.78% |
| **Myelitis transverse** | **81** | **0.46%** | **0** | **0.00%** | **3** | **0.08%** | **15** | **0.33%** | **5** | **0.29%** | **2** | **0.14%** | **1** | **0.08%** | **0** | **0.00%** | **107** | **0.34%** |
| F | 55 | 67.90% | 0 | 0.00% | 1 | 33.33% | 8 | 53.33% | 5 | 100.00% | 1 | 50.00% | 1 | 100.00% | 0 | 0.00% | 71 | 66.36% |
| M | 26 | 32.10% | 0 | 0.00% | 2 | 66.67% | 7 | 46.67% | 0 | 0.00% | 1 | 50.00% | 0 | 0.00% | 0 | 0.00% | 36 | 33.64% |
| **Myocardial infarction** | **1292** | **7.29%** | **4** | **1.02%** | **39** | **1.07%** | **174** | **3.86%** | **139** | **7.97%** | **156** | **10.65%** | **138** | **10.47%** | **111** | **15.52%** | **2053** | **6.52%** |
| F | 487 | 37.69% | 1 | 25.00% | 17 | 43.59% | 54 | 31.03% | 37 | 26.62% | 63 | 40.38% | 69 | 50.00% | 67 | 60.36% | 795 | 38.72% |
| M | 805 | 62.31% | 3 | 75.00% | 22 | 56.41% | 120 | 68.97% | 102 | 73.38% | 93 | 59.62% | 69 | 50.00% | 44 | 39.64% | 1258 | 61.28% |
| **Myocarditis or Pericarditis** | **4975** | **28.05%** | **240** | **60.91%** | **1584** | **43.55%** | **845** | **18.76%** | **235** | **13.48%** | **149** | **10.17%** | **86** | **6.53%** | **28** | **3.92%** | **8142** | **25.84%** |
| F | 1810 | 36.38% | 27 | 11.25% | 367 | 23.17% | 362 | 42.84% | 108 | 45.96% | 63 | 42.28% | 47 | 54.65% | 18 | 64.29% | 2802 | 34.41% |
| M | 3165 | 63.62% | 213 | 88.75% | 1217 | 76.83% | 483 | 57.16% | 127 | 54.04% | 86 | 57.72% | 39 | 45.35% | 10 | 35.71% | 5340 | 65.59% |
| **Narcolepsy** | **29** | **0.16%** | **1** | **0.25%** | **5** | **0.14%** | **8** | **0.18%** | **1** | **0.06%** | **1** | **0.07%** | **3** | **0.23%** | **1** | **0.14%** | **49** | **0.16%** |
| F | 20 | 68.97% | 0 | 0.00% | 5 | 100.00% | 4 | 50.00% | 1 | 100.00% | 1 | 100.00% | 1 | 33.33% | 1 | 100.00% | 33 | 67.35% |
| M | 9 | 31.03% | 1 | 100.00% | 0 | 0.00% | 4 | 50.00% | 0 | 0.00% | 0 | 0.00% | 2 | 66.67% | 0 | 0.00% | 16 | 32.65% |
| **Non-haemorrhagic or Haemorrhagic stroke** | **1880** | **10.60%** | **6** | **1.52%** | **27** | **0.74%** | **186** | **4.13%** | **182** | **10.44%** | **272** | **18.57%** | **344** | **26.10%** | **256** | **35.80%** | **3153** | **10.01%** |
| F | 956 | 50.85% | 2 | 33.33% | 18 | 66.67% | 86 | 46.24% | 65 | 35.71% | 122 | 44.85% | 183 | 53.20% | 169 | 66.02% | 1601 | 50.78% |
| M | 924 | 49.15% | 4 | 66.67% | 9 | 33.33% | 100 | 53.76% | 117 | 64.29% | 150 | 55.15% | 161 | 46.80% | 87 | 33.98% | 1552 | 49.22% |
| **Pulmonary embolism** | **3473** | **19.58%** | **5** | **1.27%** | **168** | **4.62%** | **387** | **8.59%** | **263** | **15.09%** | **257** | **17.54%** | **276** | **20.94%** | **118** | **16.50%** | **4947** | **15.70%** |
| F | 1769 | 50.94% | 3 | 60.00% | 104 | 61.90% | 169 | 43.67% | 86 | 32.70% | 112 | 43.58% | 157 | 56.88% | 75 | 63.56% | 2475 | 50.03% |
| M | 1704 | 49.06% | 2 | 40.00% | 64 | 38.10% | 218 | 56.33% | 177 | 67.30% | 145 | 56.42% | 119 | 43.12% | 43 | 36.44% | 2472 | 49.97% |
| **Grand Total** | **17735** | **56.28%** | **394** | **1.25%** | **3637** | **11.54%** | **4504** | **14.29%** | **1743** | **5.53%** | **1465** | **4.65%** | **1318** | **4.18%** | **715** | **2.27%** | **31511** | **100.00%** |

## Discussion

Vaccination saves 2.5 million children from death each year, according to WHO estimates ^21–24^. Moreover, it is the most effective technique for preventing infectious diseases and reducing their associated mortality and morbidity rate ^21,23,24^. However, threatening factors associated with vaccinations, such as AEFIs, could have a negative impact on vaccination rates ^21,23^. Here, we describe VAERS data and evaluate their usability. We also examine COVID19’s impact on the reporting patterns.

The VAERS provides a wealth of information for analyzing AEFIs and their patterns over time ^10,12,25^. Such a surveillance system with a large data set can be utilized for event detection and generation to investigate and evaluate vaccine safety, ultimately helping develop regulatory laws and actions to minimize the risks associated with these vaccines ^10,25–27^. Descriptive and pattern analysis of raw VAERS data combined with data visualization showed several intriguing patterns over the previous 31 years. This study’s outcomes provide some insights into and can guide further research on certain vaccines and AEFIs.

The COVID-19 vaccine was released under the EUA, which requires the reporting of any AEFI occurrence ^13,15^. The VAERS data shows a huge increase in reporting in 2021, accounting for 48.52% of domestic and 70.33% of nondomestic reports over the past 31 years. Furthermore, COVID-19 vaccines were the most frequently reported vaccines, accounting for 43.72% of domestic and 69.16% of nondomestic total reported vaccines.

A major component of the VAERS design is the identification of serious AEFIs, such as death, hospitalization, lethal threat, and permanent disability ^28^. The reporting rate of domestic deaths steadily decreased for 30 years until 2021, where it started rising. However, underreporting clearly occurred, with reported deaths representing only 1% of the data. We suspect underreporting in the nondomestic data as well, with reported deaths representing only 4.08% of the data. A continuous decrease in reported death occurred in the nondomestic data until 2000. The fact that the pneumococcal conjugate vaccine was released in 2000 may explain this increase in reported deaths, which also occurred in 2021, when the COVID-19 vaccine was released ^13,14,29–31^. The number of Pnumovax-associated deaths increased in 2000, confirming our suggestion ^29–31^. Another pattern worth noting is that reported domestic deaths associated with influenza vaccines increased in 2011. We found a similar increase in 2019. Influenza H1N1 was a pandemic disease that struck between 2009 and 2010; as a result, the CDC updated its 2010-2011 vaccination recommendations ^32–34^. The CDC recommended administering influenza vaccines to all persons 6 months or older ^35,36^. In 2019, the US health strategy to manage the COVID-19 pandemic included the influenza vaccination to minimize influenza-related respiratory diseases and optimize health care services for COVID-19 patients ^37^. Furthermore, nondomestic reported deaths show an interesting pattern associated with DTP, whose occurrence sudden increased in 2018, possibly due to an increase in pertussis cases in 2018 and 2019, as the CDC reported more than 15 600 cases per year ^38,39^.

Domestic ER or doctor office visit reports show an upward trend from 1990 to 2007, then show a downward pattern until they flatten. However, for nondomestic ER or doctor office visits, the pattern fluctuated steadily until 2011, when it began to rise, reaching its peak in 2015. Among the vaccines most frequently associated with nondomestic reported ER or doctor visits, the HPV vaccine showed a similar pattern. In addition, a similar increase in domestic reported ER or doctor visits was associated with HPV from 2006 to 2008, with a steady decline following. The USFDA first approved the HPV vaccine in 2006, which may explain the increase in domestic ER or doctor visits ^40^. In 2011, the Advisory Committee on Immunization Practices (ACIP) updated the recommendations for HPV vaccination to include boys, which could have led to an increase in vaccinations and associated ER visits ^41–44^.

Over the last 31 years, domestic and nondomestic hospitalization reports have steadily declined. However, supporting our previous interpretations, domestic data on the HPV vaccine showed an increased rate of hospitalization from 2006 to 2015 ^41–44^. Furthermore, the influenza vaccine was associated with a similar increase in 2011, followed by a steady decline. In addition, the USFDA approved a new Zoster (Shingrix) vaccine in 2017, which could explain the increase in hospitalizations rate associated with the Zoster in the same year ^45^. However, we found no clear cause for the sudden increase in 2015.

Reports of domestic lethal threats increased in 1998. A similar trend occurred with the hepatitis B vaccine. This increase may have occurred due to the ACIP’s recommendation in 1998 to expand the children vaccination program to include the hepatitis B vaccine ^46^. However, we found a similar increase in reports of nondomestic lethal threats in 1991, when the ACIP first recommended hepatitis B vaccines for all infants ^47^.

Domestic disabilities showed a similar trend as lethal threats associated with hepatitis B vaccines in 1991 and 1998 ^46–48^. We found another interesting increase in 1999 and a similar pattern associated with the Lyme vaccine, which the FDA approved in 1998 but its manufacturer has since withdrawn ^49,50^. We also found a similar pattern of Zoster-associated disabilities and lethal threat pattern from 2006 to 2015^45^. In 2005, we found an increase in the administration of the anthrax vaccine, which could have occurred due to the EUA’s extension of the anthrax-adsorbed vaccine, which the USFDA announced that year ^51^. Moreover, nondomestic reported disabilities show a similar pattern associated with the Zoster vaccine ^45^. We found another interesting pattern described by an increase in nondomestic disabilities in 1998, which was associated with a similar pattern in hepatitis B and MMR II vaccines. Wakefield study results may affect the reporting pattern of MMR. The outcome of this study, which was published in 1998, showed that a behavioral and development disorder in children resulted from MMR ^52,53^.

In the domestic and nondomestic data, pyrexia was the most frequently reported AEFI over the past 30 years. Pyrexia is defined as an elevation of body temperature that exceeds the normal range due to the presence of a foreign substance in the body ^54^. This AEFI is expected as a part of the activated immune response ^3,54–56^. However, in 2021, headache was the most frequently reported AEFI and was mainly associated with COVID-19 vaccines ^3,55–58^. In the domestic data, injection site AEFIs were the most common ^3,55^. Furthermore, the nondomestic data shows a variety of symptoms, with vomiting being the commonly reported AEFI over the past 30 years ^3,55,56^. CDC estimated 0.48 cases of myocarditis per 100,000 after COVID-19 vaccination, and in this study results of AESIs analysis show that most most reported adverse event associated with COVID-19 was myocarditis or pericarditis which support these results^59,60^.

Results of our study show that most of AEFIs are nonserious and more than 80% of them appear within first few days following vaccination, which is consistent with results found in other articles ^61–63^. In addition, male and age group ≤ 5 years were more prone to serious AEFIs in consistent with findings from prior research ^63,64^. These findings could strengthen the argument for VARES’s efficient use.

Generally, stimulated reporting is noticeable in this study’s outcomes ^25,26^. Yearly patterns were mostly affected by the USFDA’s approval of vaccines such as the Lyme and HPV vaccines, CDC recommendations, such as administration of the H1N1 and DTP vaccines, and ACIP recommendations, such as administration of the HPV and HEB B vaccines. In addition, pandemic diseases, such as COVID-19 and H1N1, have a major impact on vaccination pattern ^29–51,57^. Finally, public research outcomes, such as in the case of the MMR vaccine, have a noticeable impact on vaccination rate ^52,53^.

Given that this is a passive reporting system, and that each report may contain information about several vaccines, we cannot confirm a causal relationship between a vaccine and its associated AEFIs ^12,25,26^. Incomplete reports and underreporting are a noticeable issue in these data sets, which can be seen clearly with serious AEFIs. In addition, most of the nondomestic data are reported by manufacturers to fulfill legal requirements, which could lead to misleading selective reporting and thereby impact nondomestic data more than domestic data ^25^. Although the VAERS uses MedDRA medical terminology, spelling mistakes and duplication (e.g., pyrexia and elevated body temperature) could have a negative impact on the data ^12^.

In conclusion, the VAERS is considered an important and useful tool in vaccine safety surveillance. However, the usability of VAERS data depends on users understating this surveillance system’s limitations and knowing how to interpret its results. Governmental regulations, availability of vaccines, and public health recommendations have the largest impact on reporting rates, which could be used to improve VAERS data.

## Supporting information

Supplementary Figures (charts)

## Data Availability

All data produced are available online at https://vaers.hhs.gov/data.html

https://vaers.hhs.gov/data.html

## References

1. Pharmacovigilance V. Definition and Application of Terms for Vaccine Pharmacovigilance This report from the Council for International Organizations of Medical Sciences (CIOMS) in collaboration with WHO covers the activities and outputs of the CIOMS/WHO Working Group on. Published online 2005. Accessed February 6, 2022. http://www.who.int/bookorders

2. Vaccine Testing and Approval Process | CDC. Accessed February 11, 2022. https://www.cdc.gov/vaccines/basics/test-approve.html

3. Gershwin LJ. Adverse Reactions to Vaccination. Veterinary Clinics of North America: Small Animal Practice. 2018;48(2):279–290. doi:10.1016/j.cvsm.2017.10.005

4. Vaccine and Related Biological Product Guidances | FDA. Accessed February 11, 2022. https://www.fda.gov/vaccines-blood-biologics/biologics-guidances/vaccine-and-related-biological-product-guidances

5. Definition and Application of Terms for Vaccine Pharmacovigilance. Report of CIOMS/WHO Working Group on Vaccine Pharmacovigilance. .; 2012.

6. Milstien JB. Regulation of Vaccines: Strengthening the Science Base.

7. CFR - Code of Federal Regulations Title 21.

8. Food and Drug Administration Regulation and Evaluation of Vaccines abstract. Published online 2011. doi:10.1542/peds.2010-1722E

9. MODULE 1 – Pre-licensure vaccine safety - WHO Vaccine Safety Basics. Accessed February 6, 2022. https://vaccine-safety-training.org/pre-licensure-vaccine-safety.html

10. Ren JJ, Sun T, He Y, Zhang Y. A statistical analysis of vaccine-adverse event data. doi:10.1186/s12911-019-0818-8

11. ACIP Meetings Information | CDC. Accessed February 6, 2022. https://www.cdc.gov/vaccines/acip/meetings/index.html

12. VAERS - About Us. Accessed February 6, 2022. https://vaers.hhs.gov/about.html

13. Fda, Cber. Contains Nonbinding Recommendations Emergency Use Authorization for Vaccines to Prevent COVID-19 Guidance for Industry Preface Public Comment. Published online 2021. Accessed February 6, 2022. https://www.fda.gov/regulatory-

14. Hause AM, Gee J, Baggs J, et al. COVID-19 Vaccine Safety in Adolescents Aged 12–17 Years — United States, December 14, 2020–July 16, 2021.; 2020. https://www.cdc.gov/mmwr/mmwr_continuingEducation.html

15. Emergency Use Authorization for Vaccines Explained | FDA. Accessed February 11, 2022. https://www.fda.gov/vaccines-blood-biologics/vaccines/emergency-use-authorization-vaccines-explained

16. CBER Surveillance Program Background Rates of Adverse Events of Special Interest for COVID-19 Vaccine Safety Monitoring Protocol. Published online 2021. Accessed March 23, 2022. https://www.bestinitiative.org/wp-

17. Tsz Tsun Lai F, Huang L, Sze Ling Chui C, et al. Multimorbidity and adverse events of special interest associated with Covid-19 vaccines in Hong Kong. doi:10.1038/s41467-022-28068-3

18. Li X, Ostropolets A, Makadia R, et al. Characterising the background incidence rates of adverse events of special interest for covid-19 vaccines in eight countries: multinational network cohort study. doi:10.1136/bmj.n1435

19. Kartsonaki C. Survival analysis. Diagnostic Histopathology. 2016;22(7):263–270. doi:10.1016/J.MPDHP.2016.06.005

20. Kochhar S, Excler JL, Bok K, et al. Defining the interval for monitoring potential adverse events following immunization (AEFIs) after receipt of live viral vectored vaccines. Vaccine. 2019;37(38):5796–5802. doi:10.1016/J.VACCINE.2018.08.085

21. Duclos P, Okwo-Bele JM, Gacic-Dobo M, Cherian T. BMC International Health and Human Rights Opinion Global immunization: status, progress, challenges and future. doi:10.1186/1472-698X-9-S1-S2

22. Centers for Disease Control and Prevention (CDC). Ten great public health achievements-nited States, 1900-1999. MMWR Morbidity and mortality weekly report. 1999;48(12):241–243.

23. Dubé E, Laberge C, Guay M, Bramadat P, Roy R, Bettinger JA. Human Vaccines & Immunotherapeutics Vaccine hesitancy An overview. 2013;(8):1763–1773. doi:10.4161/hv.24657

24. Canouï E, Launay O. [History and principles of vaccination]. Revue des maladies respiratoires. 2019;36(1):74–81. doi:10.1016/J.RMR.2018.02.015

25. Shimabukuro TT, Nguyen M, Martin D, DeStefano F. Safety monitoring in the Vaccine Adverse Event Reporting System (VAERS). Vaccine. 2015;33(36):4398–4405. doi:10.1016/J.VACCINE.2015.07.035

26. Varricchio F, Iskander J, Destefano F, et al. Understanding vaccine safety information from the Vaccine Adverse Event Reporting System. The Pediatric infectious disease journal. 2004;23(4):287–294. doi:10.1097/00006454-200404000-00002

27. CDC, Ncird. Understanding the Vaccine Adverse Event Reporting System (VAERS). Accessed February 11, 2022. http://www.cdc.gov/vaccines/conversations

28. What is a Serious Adverse Event? | FDA. Accessed February 6, 2022. https://www.fda.gov/safety/reporting-serious-problems-fda/what-serious-adverse-event

29. DeStefano F. Safety profile of pneumococcal conjugate vaccines: systematic review of pre-and post-licensure data. Bulletin of the World Health Organization. 2008;86(5):373–380. doi:10.2471/BLT.07.048025

30. Jefferies JM, Macdonald E, Faust SN, Clarke SC. Human Vaccines 13-valent pneumococcal conjugate vaccine (PCV13). Landes Bioscience 1012 Human Vaccines. 2011;7(10):10. doi:10.4161/hv.7.10.16794

31. Pinkbook: Pneumococcal Disease | CDC. Accessed February 11, 2022. https://www.cdc.gov/vaccines/pubs/pinkbook/pneumo.html

32. Jain S, Kamimoto L, Bramley AM, et al. Hospitalized Patients with 2009 H1N1 Influenza in the United States, April–June 2009. New England Journal of Medicine. 2009;361(20):1935–1944. doi:10.1056/NEJMOA0906695/SUPPL_FILE/NEJM_JAIN_1935SA1.PDF

33. Clinical Aspects of Pandemic 2009 Influenza A (H1N1) Virus Infection. New England Journal of Medicine. 2010;362(18):1708–1719. doi:10.1056/NEJMRA1000449/SUPPL_FILE/NEJM_WHO_CONSULTATION_ON_H1N1_1708SA1.PDF

34. 2009 H1N1 Pandemic (H1N1pdm09 virus) | Pandemic Influenza (Flu) | CDC. Accessed February 11, 2022. https://www.cdc.gov/flu/pandemic-resources/2009-h1n1-pandemic.html

35. Fiore AE, Uyeki TM, Broder K, et al. Prevention and control of influenza with vaccines: recommendations of the Advisory Committee on Immunization Practices (ACIP), 2010. MMWR Recommendations and reports : Morbidity and mortality weekly report Recommendations and reports. 2010;59(RR-8):1–62.

36. Campbell AJP, Grohskopf LA. Updates on Influenza Vaccination in Children. Infectious Disease Clinics of North America. 2018;32(1):75–89. doi:10.1016/j.idc.2017.11.005

37. Grohskopf LA, Alyanak E, Ferdinands JM, et al. Prevention and Control of Seasonal Influenza with Vaccines: Recommendations of the Advisory Committee on Immunization Practices, United States, 2021–22 Influenza Season. MMWR Recommendations and Reports. 2021;70(5):1–32. doi:10.15585/MMWR.RR7005A1

38. About Diphtheria, Tetanus, and Pertussis Vaccination | CDC. Accessed February 11, 2022. https://www.cdc.gov/vaccines/vpd/dtap-tdap-td/hcp/about-vaccine.html

39. Ask the Experts about Pertussis Vaccines (DTaP, Tdap) - CDC experts answer Q&As. Accessed February 11, 2022. https://www.immunize.org/askexperts/experts_per.asp

40. Gardasil Vaccine Safety | FDA. Accessed February 11, 2022. https://www.fda.gov/vaccines-blood-biologics/safety-availability-biologics/gardasil-vaccine-safety

41. Recommendations on the Use of Quadrivalent Human Papillomavirus Vaccine in Males — Advisory Committee on Immunization Practices (ACIP), 2011. Accessed February 11, 2022. https://www.cdc.gov/mmwr/preview/mmwrhtml/mm6050a3.htm

42. Quadrivalent Human Papillomavirus Vaccine Recommendations of the Advisory Committee on Immunization Practices (ACIP). Accessed February 11, 2022. https://www.cdc.gov/mmwr/preview/mmwrhtml/rr5602a1.htm

43. Markowitz LE, Tsu V, Deeks SL, et al. Human Papillomavirus Vaccine Introduction – The First Five Years. Vaccine. 2012;30(SUPPL.5):F139–F148. doi:10.1016/J.VACCINE.2012.05.039

44. Hawkins SS, Horvath K, Cohen J, Pace LE, Baum CF. Associations between ACA-related policies and a clinical recommendation with HPV vaccine initiation. Cancer causes & control : CCC. 2021;32(7):783–790. doi:10.1007/S10552-021-01430-4

45. Shah RA, Limmer AL, Nwannunu CE, Patel RR, Mui UN, Tyring SK. Shingrix for Herpes Zoster: A Review. Skin therapy letter. 2019;24(4):5–7. Accessed February 11, 2022. https://pubmed.ncbi.nlm.nih.gov/31339679/

46. Zhao Z, Murphy T v., Jacques-Carroll L. Progress in newborn hepatitis B vaccination by birth year cohorts-1998-2007, USA. Vaccine. 2011;30(1):14–20. doi:10.1016/J.VACCINE.2011.10.076

47. Freed GL, Bordley WC, Clark SJ, Konrad TR. Universal Hepatitis B Immunization of Infants: Reactions of Pediatricians and Family Physicians Over Time. Pediatrics. 1994;93(5):747–751. doi:10.1542/PEDS.93.5.747

48. Schillie S, Murphy T v., Sawyer M, et al. CDC guidance for evaluating health-care personnel for hepatitis B virus protection and for administering postexposure management. MMWR Recommendations and reports : Morbidity and mortality weekly report Recommendations and reports. 2013;62(RR-10). Accessed February 11, 2022. https://pubmed.ncbi.nlm.nih.gov/24352112/

49. Shen AK, Mead PS, Beard CB. The Lyme Disease Vaccine—A Public Health Perspective. Clinical Infectious Diseases. 2011;52(suppl_3):s247–s252. doi:10.1093/cid/ciq115

50. Nigrovic LE, Thompson KM. The Lyme vaccine: a cautionary tale. Epidemiology and Infection. 2007;135(1):1–8. doi:10.1017/S0950268806007096

51. Federal Register :: Authorization of Emergency Use of Anthrax Vaccine Adsorbed for Prevention of Inhalation Anthrax by Individuals at Heightened Risk of Exposure Due to Attack With Anthrax; Extension; Availability. Accessed February 11, 2022. https://www.federalregister.gov/documents/2005/08/03/05-15233/authorization-of-emergency-use-of-anthrax-vaccine-adsorbed-for-prevention-of-inhalation-anthrax-by

52. Sathyanarayana Rao T, Andrade C. The MMR vaccine and autism: Sensation, refutation, retraction, and fraud. Indian Journal of Psychiatry. 2011;53(2):95. doi:10.4103/0019-5545.82529

53. Godlee F, Smith J, Marcovitch H. Wakefield’s article linking MMR vaccine and autism was fraudulent. BMJ. 2011;342(jan05 1):c7452–c7452. doi:10.1136/bmj.c7452

54. Kohl KS, Marcy SM, Blum M, et al. Fever after immunization: Current concepts and improved future scientific understanding. Clinical Infectious Diseases. 2004;39(3):389–394. doi:10.1086/422454/2/39-3-389-TBL001.GIF

55. Hervé C, Laupèze B, Giudice G del, Didierlaurent AM, Tavares F, Silva D. The how’s and what’s of vaccine reactogenicity. doi:10.1038/s41541-019-0132-6

56. Spencer JP, Trondsen Pawlowski RH, Thomas S. Vaccine Adverse Events: Separating Myth from Reality. American family physician. 2017;95(12):786–794.

57. Zhu FC, Li YH, Guan XH, et al. Safety, tolerability, and immunogenicity of a recombinant adenovirus type-5 vectored COVID-19 vaccine: a dose-escalation, open-label, non-randomised, first-in-human trial. Lancet (London, England). 2020;395(10240):1845–1854. doi:10.1016/S0140-6736(20)31208-3

58. Cocores A, Monteith T. Post-Vaccination Headache Reporting Trends According to the Vaccine Adverse Events Reporting System (VAERS) (P1.147). Neurology. 2016;86(16 Supplement):P1.147. http://n.neurology.org/content/86/16_Supplement/P1.147.abstract

59. Wallace M, Oliver S, Meeting A. http://cdc.gov/coronavirus COVID-19 mRNA vaccines in adolescents and young adults: Benefit-risk discussion. xPublished online 2021.

60. Witberg G, Barda N, Hoss S, et al. Myocarditis after Covid-19 Vaccination in a Large Health Care Organization. New England Journal of Medicine. 2021;385(23):2132–2139. doi:10.1056/NEJMoa2110737

61. Alguacil-Ramos AM, Muelas-Tirado J, Garrigues-Pelufo TM, et al. Surveillance for adverse events following immunization (AEFI) for 7 years using a computerised vaccination system. Public Health. 2016;135:66–74. doi:10.1016/j.puhe.2015.11.010

62. Mentzer D, Oberle D, Keller-Stanislawski B. Adverse events following immunisation with a meningococcal serogroup B vaccine: report from post-marketing surveillance, Germany, 2013 to 2016. Eurosurveillance. 2018;23(17). doi:10.2807/1560-7917.ES.2018.23.17.17-00468

63. Fadare JO, Haazen L, Lombardi N, et al. Vaccines Safety in Children and in General Population: A Pharmacovigilance Study on Adverse Events Following Anti-Infective Vaccination in Italy. 2019;10:948. doi:10.3389/fphar.2019.00948

64. Harris T, Nair J, Fediurek J, Deeks SL. Assessment of sex-specific differences in adverse events following immunization reporting in Ontario, 2012–15. Vaccine. 2017;35(19):2600–2604. doi:10.1016/J.VACCINE.2017.03.035

