## Supplementary Figures (charts) for "Vaccine Adverse Event Reporting System (VAERS): Evaluation of 31 Years of Reports and Pandemics’ Impact"

**Figure 1:** Number of domestic reports per year

|  | Reports |
| --- | --- |
| 1990 | 2102 |
| 1991 | 9933 |
| 1992 | 10692 |
| 1993 | 10147 |
| 1994 | 10193 |
| 1995 | 10001 |
| 1996 | 10771 |
| 1997 | 11006 |
| 1998 | 9949 |
| 1999 | 12123 |
| 2000 | 14105 |
| 2001 | 13359 |
| 2002 | 14074 |
| 2003 | 16757 |
| 2004 | 15324 |
| 2005 | 15581 |
| 2006 | 17313 |
| 2007 | 28226 |
| 2008 | 29766 |
| 2009 | 32786 |
| 2010 | 31582 |
| 2011 | 25408 |
| 2012 | 26668 |
| 2013 | 29736 |
| 2014 | 34340 |
| 2015 | 44422 |
| 2016 | 45706 |
| 2017 | 38911 |
| 2018 | 49137 |
| 2019 | 48443 |
| 2020 | 50205 |
| 2021 | 677514 |

**Figure 2:** Pattern of domestic reported death over years

|  | Death% |
| --- | --- |
| 1990 | 3.6156% |
| 1991 | 2.61754% |
| 1992 | 2.03891% |
| 1993 | 2.15827% |
| 1994 | 2.18778% |
| 1995 | 1.39986% |
| 1996 | 1.14196% |
| 1997 | 1.20843% |
| 1998 | 1.32677% |
| 1999 | 1.18782% |
| 2000 | 0.999646% |
| 2001 | 1.30249% |
| 2002 | 0.959216% |
| 2003 | 1.18756% |
| 2004 | 1.05717% |
| 2005 | 0.840768% |
| 2006 | 0.710449% |
| 2007 | 0.573939% |
| 2008 | 0.611436% |
| 2009 | 0.582566% |
| 2010 | 0.509784% |
| 2011 | 0.680888% |
| 2012 | 0.622469% |
| 2013 | 0.433818% |
| 2014 | 0.381479% |
| 2015 | 0.335419% |
| 2016 | 0.387258% |
| 2017 | 0.331526% |
| 2018 | 0.335796% |
| 2019 | 0.377764% |
| 2020 | 0.33662% |
| 2021 | 1.31746% |

**Figure 3:** Patterns over years of top 5 vaccines in domestic reported deaths

|  | INFLUENZA (SEASONAL) (NO BRAND NAME) | INFLUENZA (SEASONAL) (FLUZONE) | DTAP + HEPB + IPV (PEDIARIX) | DTP (NO BRAND NAME) | DTP (TRI-IMMUNOL) |
| --- | --- | --- | --- | --- | --- |
| 1990 | 11 | 6 | 0 | 36 | 93 |
| 1991 | 31 | 14 | 0 | 70 | 190 |
| 1992 | 94 | 28 | 0 | 146 | 334 |
| 1993 | 73 | 43 | 0 | 315 | 169 |
| 1994 | 34 | 38 | 0 | 203 | 67 |
| 1995 | 0 | 38 | 0 | 114 | 28 |
| 1996 | 10 | 17 | 0 | 80 | 19 |
| 1997 | 19 | 29 | 0 | 37 | 12 |
| 1998 | 19 | 12 | 0 | 47 | 0 |
| 1999 | 7 | 39 | 0 | 16 | 6 |
| 2000 | 4 | 18 | 0 | 10 | 1 |
| 2001 | 47 | 42 | 0 | 0 | 11 |
| 2002 | 37 | 22 | 0 | 5 | 0 |
| 2003 | 15 | 214 | 14 | 7 | 0 |
| 2004 | 34 | 252 | 124 | 1 | 0 |
| 2005 | 51 | 114 | 119 | 5 | 0 |
| 2006 | 31 | 50 | 78 | 0 | 0 |
| 2007 | 22 | 26 | 133 | 0 | 0 |
| 2008 | 18 | 51 | 152 | 0 | 0 |
| 2009 | 40 | 49 | 55 | 0 | 0 |
| 2010 | 71 | 73 | 24 | 0 | 0 |
| 2011 | 210 | 240 | 32 | 22 | 0 |
| 2012 | 61 | 66 | 58 | 9 | 0 |
| 2013 | 116 | 32 | 39 | 0 | 0 |
| 2014 | 27 | 51 | 73 | 0 | 0 |
| 2015 | 69 | 17 | 79 | 30 | 0 |
| 2016 | 123 | 23 | 120 | 6 | 0 |
| 2017 | 51 | 0 | 79 | 0 | 0 |
| 2018 | 121 | 0 | 63 | 6 | 0 |
| 2019 | 209 | 2 | 43 | 0 | 0 |
| 2020 | 118 | 0 | 88 | 0 | 0 |
| 2021 | 17 | 0 | 8 | 0 | 0 |

**Figure 4**: Pattern of domestic ER or doctor visits over years

|  | ER% |
| --- | --- |
| 1990 | 16.4605% |
| 1991 | 32.4071% |
| 1992 | 35.9147% |
| 1993 | 40.2385% |
| 1994 | 40.3218% |
| 1995 | 42.0658% |
| 1996 | 36.1619% |
| 1997 | 35.417% |
| 1998 | 35.3402% |
| 1999 | 38.9095% |
| 2000 | 43.3534% |
| 2001 | 44.4419% |
| 2002 | 44.3015% |
| 2003 | 42.0421% |
| 2004 | 40.7204% |
| 2005 | 41.8779% |
| 2006 | 46.2774% |
| 2007 | 47.3074% |
| 2008 | 38.8363% |
| 2009 | 35.5792% |
| 2010 | 33.6552% |
| 2011 | 34.7607% |
| 2012 | 32.4246% |
| 2013 | 28.9582% |
| 2014 | 24.7612% |
| 2015 | 19.6929% |
| 2016 | 19.4701% |
| 2017 | 9.94577% |
| 2018 | 1.72986% |
| 2019 | 0.641992% |
| 2020 | 0.346579% |
| 2021 | 0.00782272% |

**Figure 5:** Patterns over years of top 5 vaccines in domestic reported ER or doctor visits

|  | INFLUENZA (SEASONAL) (FLUZONE) | HPV (GARDASIL) | PNEUMO (PNEUMOVAX) | VARICELLA (VARIVAX) | MEASLES + MUMPS + RUBELLA (MMR II) |
| --- | --- | --- | --- | --- | --- |
| 1990 | 92 | 0 | 2 | 0 | 93 |
| 1991 | 167 | 0 | 39 | 0 | 1137 |
| 1992 | 257 | 0 | 60 | 0 | 1526 |
| 1993 | 285 | 0 | 44 | 0 | 1050 |
| 1994 | 731 | 0 | 138 | 0 | 1177 |
| 1995 | 728 | 0 | 174 | 425 | 1211 |
| 1996 | 423 | 0 | 167 | 1070 | 1145 |
| 1997 | 756 | 0 | 180 | 1300 | 1140 |
| 1998 | 951 | 0 | 266 | 886 | 899 |
| 1999 | 1009 | 0 | 588 | 1883 | 1073 |
| 2000 | 899 | 0 | 635 | 2403 | 1435 |
| 2001 | 1046 | 0 | 723 | 2003 | 1667 |
| 2002 | 1566 | 0 | 1108 | 1574 | 1425 |
| 2003 | 1663 | 0 | 1448 | 505 | 1331 |
| 2004 | 1882 | 0 | 1604 | 2219 | 1416 |
| 2005 | 3228 | 0 | 2245 | 2211 | 1283 |
| 2006 | 2362 | 703 | 2557 | 2937 | 1703 |
| 2007 | 3340 | 9495 | 3163 | 2776 | 1266 |
| 2008 | 4015 | 9838 | 2586 | 2673 | 1298 |
| 2009 | 4195 | 6419 | 2489 | 1431 | 1225 |
| 2010 | 5246 | 4236 | 2577 | 892 | 992 |
| 2011 | 5922 | 3051 | 2321 | 930 | 1076 |
| 2012 | 4844 | 2554 | 2239 | 721 | 946 |
| 2013 | 4305 | 2902 | 2372 | 761 | 764 |
| 2014 | 3304 | 2786 | 2436 | 557 | 936 |
| 2015 | 2258 | 2510 | 2072 | 572 | 982 |
| 2016 | 777 | 1731 | 2438 | 534 | 712 |
| 2017 | 94 | 496 | 1719 | 226 | 400 |
| 2018 | 0 | 36 | 325 | 58 | 52 |
| 2019 | 0 | 16 | 102 | 22 | 31 |
| 2020 | 0 | 13 | 41 | 9 | 25 |
| 2021 | 0 | 0 | 1 | 0 | 7 |

**Figure 6:**  Pattern of domestic reported hospitalization over years

|  | Hos% |
| --- | --- |
| 1990 | 12.1313% |
| 1991 | 2.5672% |
| 1992 | 2.38496% |
| 1993 | 2.51306% |
| 1994 | 2.50172% |
| 1995 | 2.54975% |
| 1996 | 2.36747% |
| 1997 | 2.31692% |
| 1998 | 2.56307% |
| 1999 | 2.10344% |
| 2000 | 1.80787% |
| 2001 | 1.90883% |
| 2002 | 1.81185% |
| 2003 | 1.52175% |
| 2004 | 1.66406% |
| 2005 | 1.63661% |
| 2006 | 1.47288% |
| 2007 | 0.903422% |
| 2008 | 0.856682% |
| 2009 | 0.777771% |
| 2010 | 0.807422% |
| 2011 | 1.00362% |
| 2012 | 0.956202% |
| 2013 | 0.857546% |
| 2014 | 0.742574% |
| 2015 | 0.57404% |
| 2016 | 0.557914% |
| 2017 | 0.655342% |
| 2018 | 0.518957% |
| 2019 | 0.526392% |
| 2020 | 0.507918% |
| 2021 | 0.0376376% |

**Figure 7:** Patterns over years of top 5 vaccines in domestic reported hospitalization

|  | INFLUENZA (SEASONAL) (NO BRAND NAME) | INFLUENZA (SEASONAL) (FLUZONE) | HPV (GARDASIL) | PNEUMO (PNEUMOVAX) | ZOSTER (SHINGRIX) |
| --- | --- | --- | --- | --- | --- |
| 1990 | 65 | 55 | 0 | 13 | 0 |
| 1991 | 164 | 79 | 0 | 60 | 0 |
| 1992 | 384 | 144 | 0 | 33 | 0 |
| 1993 | 540 | 118 | 0 | 35 | 0 |
| 1994 | 188 | 213 | 0 | 45 | 0 |
| 1995 | 63 | 224 | 0 | 61 | 0 |
| 1996 | 79 | 152 | 0 | 63 | 0 |
| 1997 | 97 | 246 | 0 | 55 | 0 |
| 1998 | 89 | 249 | 0 | 65 | 0 |
| 1999 | 138 | 382 | 0 | 153 | 0 |
| 2000 | 78 | 157 | 0 | 106 | 0 |
| 2001 | 47 | 69 | 0 | 6 | 0 |
| 2002 | 83 | 107 | 0 | 23 | 0 |
| 2003 | 22 | 99 | 0 | 38 | 0 |
| 2004 | 163 | 110 | 0 | 50 | 0 |
| 2005 | 46 | 190 | 0 | 143 | 0 |
| 2006 | 80 | 135 | 22 | 164 | 0 |
| 2007 | 158 | 167 | 535 | 354 | 0 |
| 2008 | 91 | 331 | 1004 | 203 | 0 |
| 2009 | 388 | 172 | 1464 | 178 | 0 |
| 2010 | 148 | 197 | 838 | 309 | 0 |
| 2011 | 417 | 1942 | 550 | 387 | 0 |
| 2012 | 631 | 1261 | 806 | 606 | 0 |
| 2013 | 529 | 1185 | 766 | 547 | 0 |
| 2014 | 853 | 845 | 676 | 621 | 0 |
| 2015 | 661 | 563 | 615 | 457 | 0 |
| 2016 | 243 | 22 | 447 | 710 | 1425 |
| 2017 | 243 | 22 | 447 | 680 | 0 |
| 2018 | 1145 | 0 | 200 | 710 | 1425 |
| 2019 | 1015 | 2 | 96 | 490 | 2108 |
| 2020 | 477 | 0 | 38 | 554 | 1524 |
| 2021 | 286 | 22 | 95 | 179 | 634 |

**Figure 8:** Pattern of domestic reported lethal threat over years

|  | LT% |
| --- | --- |
| 1990 | 1.56993% |
| 1991 | 1.36917% |
| 1992 | 1.87991% |
| 1993 | 1.68523% |
| 1994 | 2.00137% |
| 1995 | 2.0298% |
| 1996 | 1.81042% |
| 1997 | 1.58096% |
| 1998 | 1.99015% |
| 1999 | 2.55712% |
| 2000 | 1.46048% |
| 2001 | 1.61689% |
| 2002 | 1.62001% |
| 2003 | 1.65304% |
| 2004 | 1.55312% |
| 2005 | 1.77781% |
| 2006 | 1.5884% |
| 2007 | 1.66159% |
| 2008 | 1.63274% |
| 2009 | 1.70195% |
| 2010 | 1.88082% |
| 2011 | 1.71599% |
| 2012 | 1.51117% |
| 2013 | 1.42252% |
| 2014 | 1.23471% |
| 2015 | 0.965738% |
| 2016 | 0.962674% |
| 2017 | 0.922618% |
| 2018 | 0.787594% |
| 2019 | 0.656442% |
| 2020 | 0.627428% |
| 2021 | 1.51864% |

**Figure 9:** Patterns over years of top 5 vaccines in domestic reported lethal threat

|  | INFLUENZA (SEASONAL) (FLUZONE) | HPV (GARDASIL) | INFLUENZA (SEASONAL) (NO BRAND NAME) | DTAP + HEPB + IPV (PEDIARIX) | INFLUENZA (SEASONAL) (FLUVIRIN) |
| --- | --- | --- | --- | --- | --- |
| 1990 | 4 | 0 | 15 | 0 | 0 |
| 1991 | 1 | 0 | 36 | 0 | 0 |
| 1992 | 38 | 0 | 94 | 0 | 0 |
| 1993 | 25 | 0 | 143 | 0 | 0 |
| 1994 | 80 | 0 | 49 | 0 | 11 |
| 1995 | 38 | 0 | 34 | 0 | 3 |
| 1996 | 57 | 0 | 38 | 0 | 17 |
| 1997 | 73 | 0 | 62 | 0 | 4 |
| 1998 | 77 | 0 | 87 | 0 | 22 |
| 1999 | 106 | 0 | 109 | 0 | 27 |
| 2000 | 86 | 0 | 18 | 0 | 15 |
| 2001 | 78 | 0 | 36 | 0 | 42 |
| 2002 | 140 | 0 | 54 | 0 | 62 |
| 2003 | 169 | 0 | 51 | 28 | 152 |
| 2004 | 273 | 0 | 122 | 60 | 87 |
| 2005 | 287 | 840 | 212 | 0 | 194 |
| 2006 | 318 | 40 | 144 | 251 | 25 |
| 2007 | 269 | 383 | 200 | 376 | 56 |
| 2008 | 299 | 491 | 59 | 266 | 133 |
| 2009 | 287 | 840 | 212 | 117 | 194 |
| 2010 | 548 | 315 | 181 | 125 | 385 |
| 2011 | 671 | 185 | 85 | 132 | 184 |
| 2012 | 514 | 215 | 63 | 128 | 210 |
| 2013 | 405 | 410 | 99 | 122 | 237 |
| 2014 | 304 | 380 | 161 | 266 | 173 |
| 2015 | 146 | 299 | 77 | 107 | 227 |
| 2016 | 120 | 318 | 207 | 188 | 102 |
| 2017 | 15 | 144 | 52 | 103 | 149 |
| 2018 | 0 | 42 | 100 | 139 | 30 |
| 2019 | 0 | 38 | 44 | 245 | 0 |
| 2020 | 0 | 2 | 37 | 84 | 0 |
| 2021 | 7 | 2 | 29 | 33 | 9 |

**Figure 10:** Pattern of domestic reported disabilities over years

|  | Dis% |
| --- | --- |
| 1990 | 3.94862% |
| 1991 | 1.69133% |
| 1992 | 1.25327% |
| 1993 | 1.6261% |
| 1994 | 2.15834% |
| 1995 | 1.49985% |
| 1996 | 1.55974% |
| 1997 | 1.60821% |
| 1998 | 1.85948% |
| 1999 | 2.34265% |
| 2000 | 2.11982% |
| 2001 | 2.81458% |
| 2002 | 2.87054% |
| 2003 | 1.74256% |
| 2004 | 1.8011% |
| 2005 | 1.68795% |
| 2006 | 1.70392% |
| 2007 | 1.78204% |
| 2008 | 1.81751% |
| 2009 | 1.62264% |
| 2010 | 1.50085% |
| 2011 | 1.36965% |
| 2012 | 1.30118% |
| 2013 | 1.15012% |
| 2014 | 1.32207% |
| 2015 | 0.997254% |
| 2016 | 1.03269% |
| 2017 | 1.47773% |
| 2018 | 1.7441% |
| 2019 | 2.14272% |
| 2020 | 1.85241% |
| 2021 | 1.60056% |

**Figure 11:** Patterns over years of top 5 vaccines in domestic reported disabilities

|  | HPV (GARDASIL) | ZOSTER LIVE (ZOSTAVAX) | INFLUENZA (SEASONAL) (FLUZONE) | MEASLES + MUMPS + RUBELLA (MMR II) | ANTHRAX (BIOTHRAX) |
| --- | --- | --- | --- | --- | --- |
| 1990 | 0 | 0 | 0 | 48 | 0 |
| 1991 | 0 | 0 | 2 | 78 | 0 |
| 1992 | 0 | 0 | 16 | 83 | 0 |
| 1993 | 0 | 0 | 8 | 48 | 5 |
| 1994 | 0 | 0 | 24 | 54 | 0 |
| 1995 | 0 | 0 | 20 | 102 | 0 |
| 1996 | 0 | 0 | 19 | 84 | 0 |
| 1997 | 0 | 0 | 27 | 71 | 0 |
| 1998 | 0 | 0 | 50 | 54 | 0 |
| 1999 | 0 | 0 | 65 | 81 | 0 |
| 2000 | 0 | 0 | 18 | 139 | 1 |
| 2001 | 0 | 0 | 69 | 192 | 83 |
| 2002 | 0 | 0 | 107 | 104 | 60 |
| 2003 | 0 | 0 | 99 | 121 | 82 |
| 2004 | 0 | 0 | 110 | 62 | 214 |
| 2005 | 0 | 0 | 190 | 201 | 275 |
| 2006 | 22 | 11 | 135 | 156 | 664 |
| 2007 | 535 | 220 | 167 | 96 | 243 |
| 2008 | 1004 | 285 | 331 | 150 | 96 |
| 2009 | 1464 | 252 | 172 | 109 | 198 |
| 2010 | 838 | 214 | 197 | 23 | 56 |
| 2011 | 264 | 157 | 208 | 51 | 84 |
| 2012 | 312 | 176 | 309 | 57 | 139 |
| 2013 | 429 | 134 | 408 | 104 | 115 |
| 2014 | 534 | 238 | 354 | 85 | 103 |
| 2015 | 695 | 131 | 271 | 118 | 71 |
| 2016 | 539 | 132 | 201 | 69 | 180 |
| 2017 | 342 | 488 | 36 | 214 | 98 |
| 2018 | 239 | 728 | 18 | 133 | 87 |
| 2019 | 166 | 1145 | 9 | 198 | 89 |
| 2020 | 15 | 1383 | 0 | 118 | 45 |
| 2021 | 352 | 513 | 0 | 71 | 58 |

**Figure 12:** Number of nondomestic reports per year

|  | Reports |
| --- | --- |
| 1990 | 49 |
| 1991 | 59 |
| 1992 | 125 |
| 1993 | 161 |
| 1994 | 161 |
| 1995 | 272 |
| 1996 | 418 |
| 1997 | 602 |
| 1998 | 833 |
| 1999 | 755 |
| 2000 | 1007 |
| 2001 | 1274 |
| 2002 | 1257 |
| 2003 | 1323 |
| 2004 | 1185 |
| 2005 | 1866 |
| 2006 | 2017 |
| 2007 | 2587 |
| 2008 | 3677 |
| 2009 | 4256 |
| 2010 | 4994 |
| 2011 | 5718 |
| 2012 | 5439 |
| 2013 | 6509 |
| 2014 | 6868 |
| 2015 | 7511 |
| 2016 | 7991 |
| 2017 | 7443 |
| 2018 | 8673 |
| 2019 | 9569 |
| 2020 | 8111 |
| 2021 | 243500 |

**Figure 13:** Pattern of nondomestic reported death over years

|  | Death% |
| --- | --- |
| 1990 | 8.16327% |
| 1991 | 10.1695% |
| 1992 | 8% |
| 1993 | 9.31677% |
| 1994 | 8.69565% |
| 1995 | 6.61765% |
| 1996 | 6.69856% |
| 1997 | 6.64452% |
| 1998 | 4.80192% |
| 1999 | 4.63576% |
| 2000 | 7.05065% |
| 2001 | 4.00314% |
| 2002 | 4.13683% |
| 2003 | 4.98866% |
| 2004 | 3.88186% |
| 2005 | 4.50161% |
| 2006 | 4.80912% |
| 2007 | 3.86548% |
| 2008 | 4.10661% |
| 2009 | 3.43045% |
| 2010 | 3.1638% |
| 2011 | 2.7632% |
| 2012 | 2.75786% |
| 2013 | 3.2263% |
| 2014 | 3.30518% |
| 2015 | 3.03555% |
| 2016 | 3.25366% |
| 2017 | 4.54118% |
| 2018 | 4.26611% |
| 2019 | 4.41007% |
| 2020 | 3.09456% |
| 2021 | 4.22136% |

**Figure 14:** Patterns over years of top 5 vaccines in nondomestic reported death

|  | INFLUENZA (SEASONAL) (NO BRAND NAME) | PNEUMO (PNEUMOVAX) | PNEUMO (PREVNAR13) | DTP (NO BRAND NAME) | HPV (GARDASIL) |
| --- | --- | --- | --- | --- | --- |
| 1990 | 0 | 0 | 0 | 0 | 0 |
| 1991 | 0 | 0 | 0 | 0 | 0 |
| 1992 | 0 | 1 | 0 | 1 | 0 |
| 1993 | 0 | 3 | 0 | 2 | 0 |
| 1994 | 0 | 2 | 0 | 0 | 0 |
| 1995 | 3 | 0 | 0 | 3 | 0 |
| 1996 | 0 | 2 | 0 | 8 | 0 |
| 1997 | 1 | 1 | 0 | 12 | 0 |
| 1998 | 0 | 2 | 0 | 5 | 0 |
| 1999 | 3 | 2 | 0 | 6 | 0 |
| 2000 | 4 | 16 | 0 | 8 | 0 |
| 2001 | 2 | 4 | 0 | 5 | 0 |
| 2002 | 1 | 3 | 0 | 3 | 0 |
| 2003 | 2 | 3 | 0 | 0 | 0 |
| 2004 | 2 | 9 | 0 | 2 | 0 |
| 2005 | 6 | 12 | 0 | 2 | 0 |
| 2006 | 8 | 26 | 0 | 4 | 0 |
| 2007 | 0 | 9 | 0 | 2 | 2 |
| 2008 | 6 | 8 | 0 | 5 | 3 |
| 2009 | 19 | 12 | 0 | 3 | 9 |
| 2010 | 14 | 12 | 1 | 3 | 12 |
| 2011 | 14 | 7 | 14 | 9 | 5 |
| 2012 | 30 | 14 | 8 | 5 | 6 |
| 2013 | 34 | 7 | 29 | 6 | 10 |
| 2014 | 16 | 8 | 23 | 4 | 44 |
| 2015 | 15 | 12 | 26 | 3 | 21 |
| 2016 | 14 | 18 | 27 | 4 | 13 |
| 2017 | 31 | 13 | 22 | 13 | 19 |
| 2018 | 24 | 9 | 37 | 68 | 26 |
| 2019 | 39 | 10 | 22 | 1 | 10 |
| 2020 | 44 | 8 | 21 | 0 | 5 |
| 2021 | 11 | 3 | 6 | 2 | 1 |

**Figure 15:** Pattern of nondomestic ER or doctor visits over years

|  | ER% |
| --- | --- |
| 1990 | 4.08163% |
| 1991 | 3.38983% |
| 1992 | 7.2% |
| 1993 | 3.10559% |
| 1994 | 4.96894% |
| 1995 | 6.25% |
| 1996 | 1.19617% |
| 1997 | 4.81728% |
| 1998 | 3.72149% |
| 1999 | 1.8543% |
| 2000 | 11.4201% |
| 2001 | 4.3956% |
| 2002 | 5.33015% |
| 2003 | 4.83749% |
| 2004 | 1.85654% |
| 2005 | 2.14362% |
| 2006 | 4.26376% |
| 2007 | 2.89911% |
| 2008 | 2.69241% |
| 2009 | 2.37312% |
| 2010 | 2.0825% |
| 2011 | 3.32284% |
| 2012 | 7.31752% |
| 2013 | 18.0673% |
| 2014 | 19.8311% |
| 2015 | 24.4974% |
| 2016 | 17.7199% |
| 2017 | 6.15343% |
| 2018 | 0.887813% |
| 2019 | 0% |
| 2020 | 0% |
| 2021 | 0% |

**Figure 16:** Patterns over years of top 5 vaccines in *non*domestic reported ER visits

|  | HPV (GARDASIL) | PNEUMO (PNEUMOVAX) | DTAP+IPV+HEPB+HIB (INFANRIX HEXA) | INFLUENZA (SEASONAL) (NO BRAND NAME) | ROTAVIRUS (ROTATEQ) |
| --- | --- | --- | --- | --- | --- |
| 1990 | 0 | 0 | 0 | 0 | 0 |
| 1991 | 0 | 0 | 0 | 0 | 0 |
| 1992 | 0 | 0 | 0 | 0 | 0 |
| 1993 | 0 | 0 | 0 | 0 | 0 |
| 1994 | 0 | 0 | 0 | 0 | 0 |
| 1995 | 0 | 0 | 0 | 0 | 0 |
| 1996 | 0 | 0 | 0 | 1 | 0 |
| 1997 | 0 | 0 | 0 | 1 | 0 |
| 1998 | 0 | 2 | 0 | 1 | 0 |
| 1999 | 0 | 0 | 0 | 0 | 0 |
| 2000 | 0 | 8 | 0 | 3 | 0 |
| 2001 | 0 | 0 | 0 | 2 | 0 |
| 2002 | 0 | 1 | 0 | 1 | 0 |
| 2003 | 0 | 2 | 0 | 0 | 0 |
| 2004 | 0 | 0 | 1 | 1 | 0 |
| 2005 | 0 | 0 | 0 | 0 | 0 |
| 2006 | 0 | 0 | 1 | 4 | 0 |
| 2007 | 0 | 0 | 2 | 0 | 1 |
| 2008 | 1 | 0 | 1 | 0 | 0 |
| 2009 | 8 | 0 | 1 | 4 | 0 |
| 2010 | 3 | 0 | 2 | 10 | 1 |
| 2011 | 14 | 2 | 6 | 10 | 2 |
| 2012 | 64 | 15 | 36 | 21 | 14 |
| 2013 | 388 | 70 | 87 | 60 | 67 |
| 2014 | 319 | 158 | 78 | 46 | 81 |
| 2015 | 586 | 197 | 91 | 58 | 107 |
| 2016 | 279 | 229 | 79 | 84 | 62 |
| 2017 | 37 | 34 | 31 | 33 | 4 |
| 2018 | 0 | 0 | 2 | 13 | 0 |

**Figure 17:** Pattern of nondomestic reported hospitalization over years

|  | Hos% |
| --- | --- |
| 1990 | 46.9388% |
| 1991 | 57.6271% |
| 1992 | 52% |
| 1993 | 72.6708% |
| 1994 | 76.3975% |
| 1995 | 66.5441% |
| 1996 | 65.0718% |
| 1997 | 68.4385% |
| 1998 | 58.3433% |
| 1999 | 54.1722% |
| 2000 | 53.9225% |
| 2001 | 45.9969% |
| 2002 | 44.8687% |
| 2003 | 40.514% |
| 2004 | 46.1603% |
| 2005 | 48.4995% |
| 2006 | 43.9266% |
| 2007 | 47.1202% |
| 2008 | 51.2918% |
| 2009 | 41.6118% |
| 2010 | 42.6512% |
| 2011 | 40.4337% |
| 2012 | 41.3863% |
| 2013 | 40.4978% |
| 2014 | 38.643% |
| 2015 | 35.8275% |
| 2016 | 35.2396% |
| 2017 | 37.5386% |
| 2018 | 35.9045% |
| 2019 | 33.0129% |
| 2020 | 34.8662% |
| 2021 | 22.676% |

**Figure 18:** Patterns over years of top 5 vaccines in nondomestic reported hospitalization

|  | HPV (GARDASIL) | PNEUMO (PNEUMOVAX) | DTAP+IPV+HEPB+HIB (INFANRIX HEXA) | HEP B (ENGERIX-B) | ROTAVIRUS (ROTARIX) |
| --- | --- | --- | --- | --- | --- |
| 1990 | 0 | 1 | 0 | 11 | 0 |
| 1991 | 0 | 1 | 0 | 16 | 0 |
| 1992 | 0 | 5 | 0 | 21 | 0 |
| 1993 | 0 | 6 | 0 | 30 | 0 |
| 1994 | 0 | 5 | 0 | 41 | 0 |
| 1995 | 0 | 3 | 0 | 74 | 0 |
| 1996 | 0 | 14 | 0 | 120 | 0 |
| 1997 | 0 | 6 | 0 | 146 | 0 |
| 1998 | 0 | 12 | 0 | 155 | 0 |
| 1999 | 0 | 12 | 0 | 123 | 0 |
| 2000 | 0 | 22 | 0 | 87 | 0 |
| 2001 | 0 | 25 | 1 | 87 | 0 |
| 2002 | 0 | 35 | 4 | 114 | 0 |
| 2003 | 0 | 39 | 6 | 85 | 0 |
| 2004 | 0 | 28 | 7 | 57 | 0 |
| 2005 | 0 | 104 | 12 | 107 | 0 |
| 2006 | 1 | 115 | 29 | 60 | 0 |
| 2007 | 145 | 124 | 21 | 63 | 0 |
| 2008 | 459 | 99 | 41 | 77 | 178 |
| 2009 | 283 | 61 | 127 | 74 | 130 |
| 2010 | 159 | 90 | 182 | 60 | 104 |
| 2011 | 166 | 59 | 191 | 57 | 88 |
| 2012 | 151 | 68 | 231 | 68 | 107 |
| 2013 | 353 | 62 | 213 | 55 | 106 |
| 2014 | 287 | 148 | 178 | 51 | 107 |
| 2015 | 459 | 178 | 172 | 62 | 177 |
| 2016 | 259 | 259 | 182 | 78 | 208 |
| 2017 | 155 | 182 | 166 | 53 | 298 |
| 2018 | 155 | 205 | 187 | 46 | 205 |
| 2019 | 97 | 139 | 150 | 53 | 195 |
| 2020 | 75 | 198 | 98 | 44 | 102 |
| 2021 | 30 | 63 | 31 | 15 | 48 |

**Figure 19:** Pattern of nondomestic reported lethal threat over years

|  | LT% |
| --- | --- |
| 1990 | 4.08163% |
| 1991 | 10.1695% |
| 1992 | 17.6% |
| 1993 | 6.8323% |
| 1994 | 9.93789% |
| 1995 | 6.98529% |
| 1996 | 5.26316% |
| 1997 | 5.81395% |
| 1998 | 5.7623% |
| 1999 | 5.69536% |
| 2000 | 6.85204% |
| 2001 | 6.20094% |
| 2002 | 6.1257% |
| 2003 | 5.44218% |
| 2004 | 6.4135% |
| 2005 | 4.34084% |
| 2006 | 4.80912% |
| 2007 | 5.29571% |
| 2008 | 4.65053% |
| 2009 | 4.06485% |
| 2010 | 3.94473% |
| 2011 | 3.42777% |
| 2012 | 3.19912% |
| 2013 | 3.16485% |
| 2014 | 2.60629% |
| 2015 | 3.44828% |
| 2016 | 3.85434% |
| 2017 | 3.4126% |
| 2018 | 3.51666% |
| 2019 | 3.03062% |
| 2020 | 3.61238% |
| 2021 | 4.61355% |

**Figure 20:** Patterns over years of top 5 vaccines in nondomestic reported lethal threat

|  | DTAP+IPV+HEPB+HIB (INFANRIX HEXA) | INFLUENZA (SEASONAL) (NO BRAND NAME) | HPV (GARDASIL) | ROTAVIRUS (ROTARIX) | HEP B (ENGERIX-B) |
| --- | --- | --- | --- | --- | --- |
| 1990 | 0 | 0 | 0 | 0 | 0 |
| 1991 | 0 | 0 | 0 | 0 | 3 |
| 1992 | 0 | 0 | 0 | 0 | 6 |
| 1993 | 0 | 0 | 0 | 0 | 3 |
| 1994 | 0 | 0 | 0 | 0 | 3 |
| 1995 | 0 | 1 | 0 | 0 | 3 |
| 1996 | 0 | 0 | 0 | 0 | 2 |
| 1997 | 0 | 1 | 0 | 0 | 9 |
| 1998 | 0 | 1 | 0 | 0 | 10 |
| 1999 | 0 | 1 | 0 | 0 | 5 |
| 2000 | 0 | 3 | 0 | 0 | 9 |
| 2001 | 1 | 1 | 0 | 0 | 8 |
| 2002 | 0 | 3 | 0 | 0 | 6 |
| 2003 | 1 | 3 | 0 | 0 | 5 |
| 2004 | 1 | 5 | 0 | 0 | 6 |
| 2005 | 3 | 13 | 0 | 0 | 5 |
| 2006 | 3 | 6 | 0 | 0 | 7 |
| 2007 | 7 | 3 | 11 | 0 | 7 |
| 2008 | 10 | 3 | 21 | 30 | 2 |
| 2009 | 16 | 8 | 14 | 10 | 25 |
| 2010 | 23 | 18 | 19 | 11 | 6 |
| 2011 | 20 | 24 | 15 | 17 | 6 |
| 2012 | 24 | 10 | 24 | 11 | 5 |
| 2013 | 20 | 17 | 25 | 16 | 6 |
| 2014 | 20 | 15 | 23 | 16 | 4 |
| 2015 | 21 | 14 | 24 | 23 | 13 |
| 2016 | 32 | 29 | 15 | 23 | 10 |
| 2017 | 14 | 22 | 16 | 19 | 9 |
| 2018 | 28 | 15 | 13 | 22 | 7 |
| 2019 | 16 | 15 | 8 | 15 | 13 |
| 2020 | 21 | 9 | 7 | 11 | 13 |
| 2021 | 5 | 32 | 3 | 3 | 3 |

**Figure 21:** Pattern of nondomestic reported disabilities over years

|  | Dis% |
| --- | --- |
| 1990 | 22.449% |
| 1991 | 6.77966% |
| 1992 | 13.6% |
| 1993 | 10.559% |
| 1994 | 14.2857% |
| 1995 | 14.3382% |
| 1996 | 13.1579% |
| 1997 | 12.4585% |
| 1998 | 14.4058% |
| 1999 | 15.7616% |
| 2000 | 13.3069% |
| 2001 | 18.2104% |
| 2002 | 16.7064% |
| 2003 | 14.5881% |
| 2004 | 10.8017% |
| 2005 | 8.68167% |
| 2006 | 6.84184% |
| 2007 | 5.95284% |
| 2008 | 6.09192% |
| 2009 | 5.85056% |
| 2010 | 5.52663% |
| 2011 | 4.17978% |
| 2012 | 4.21033% |
| 2013 | 4.79336% |
| 2014 | 6.49388% |
| 2015 | 11.1437% |
| 2016 | 7.84633% |
| 2017 | 7.64477% |
| 2018 | 6.67589% |
| 2019 | 4.79674% |
| 2020 | 5.05486% |
| 2021 | 8.31951% |

**Figure 22:** Patterns over years of top 5 vaccines in nondomestic reported disabilities

|  | HPV (GARDASIL) | HEP B (ENGERIX-B) | INFLUENZA (SEASONAL) (NO BRAND NAME) | HPV (CERVARIX) | MEASLES + MUMPS + RUBELLA (MMR II) |
| --- | --- | --- | --- | --- | --- |
| 1990 | 0 | 1 | 0 | 0 | 4 |
| 1991 | 0 | 0 | 0 | 0 | 0 |
| 1992 | 0 | 1 | 0 | 0 | 5 |
| 1993 | 0 | 2 | 0 | 0 | 4 |
| 1994 | 0 | 12 | 0 | 0 | 3 |
| 1995 | 0 | 19 | 0 | 0 | 6 |
| 1996 | 0 | 30 | 0 | 0 | 5 |
| 1997 | 0 | 39 | 3 | 0 | 7 |
| 1998 | 0 | 56 | 2 | 0 | 19 |
| 1999 | 0 | 58 | 1 | 0 | 20 |
| 2000 | 0 | 56 | 3 | 0 | 19 |
| 2001 | 0 | 78 | 2 | 0 | 36 |
| 2002 | 0 | 65 | 7 | 0 | 21 |
| 2003 | 0 | 66 | 7 | 0 | 11 |
| 2004 | 0 | 38 | 8 | 0 | 4 |
| 2005 | 0 | 57 | 7 | 0 | 6 |
| 2006 | 0 | 29 | 3 | 0 | 7 |
| 2007 | 16 | 34 | 10 | 0 | 8 |
| 2008 | 39 | 52 | 10 | 0 | 5 |
| 2009 | 40 | 31 | 19 | 15 | 9 |
| 2010 | 24 | 34 | 17 | 34 | 5 |
| 2011 | 35 | 20 | 17 | 27 | 2 |
| 2012 | 29 | 19 | 35 | 23 | 6 |
| 2013 | 76 | 26 | 38 | 39 | 4 |
| 2014 | 124 | 11 | 37 | 43 | 9 |
| 2015 | 360 | 16 | 49 | 80 | 5 |
| 2016 | 239 | 15 | 49 | 46 | 8 |
| 2017 | 127 | 18 | 36 | 44 | 7 |
| 2018 | 112 | 22 | 22 | 31 | 1 |
| 2019 | 55 | 13 | 15 | 5 | 0 |
| 2020 | 23 | 15 | 9 | 8 | 1 |
| 2021 | 13 | 9 | 72 | 4 | 1 |
